# Truncated ASXL3 Alters Chromatin Accessibility and Epigenetic Landscape in Bainbridge-Ropers Syndrome Suggesting a Gain-of-Function Etiology

**DOI:** 10.64898/2026.08.04.26359647

**Authors:** Nofar Mor, Inna Shomer, Shaul Raviv, Noga Budick-Harmelin, Tanya Matzevitch, Shalhevet Azriel, Joseph G. Gleeson, Yoach Rais, Rebecca Haffner-Krausz, Shifra Ben-Dor, Omri Nayshool, Sharon Avkin-Nachum, Adi Yahalom, Efrat Sa’ar Glick, Gali Heimer, Gideon Rechavi, Dan Dominissini

**Author notes:** Corresponding authors. Emails: Nofar Mor, Dan Dominissini. Equal Contribution (Nofar Mor, Inna Shomer).

## Abstract

Bainbridge-Ropers syndrome (BRS) is a rare neurodevelopmental disorder caused by truncating mutations in the epigenetic regulator *ASXL3*. While traditionally considered a haploinsufficiency disorder, the precise molecular mechanisms driving BRS remain poorly understood. Here, we combine patient-derived cellular lines and novel mouse models to elucidate the molecular function of disease-associated *ASXL3* variants. We show that several pathogenic *ASXL3* variants escape nonsense-mediated decay (NMD), possibly leading to the accumulation of truncated protein, and widespread epigenetic changes, resulting in distinct transcriptomic and proteomic profiles. These changes include increased chromatin accessibility and global DNA hypomethylation, particularly at promoters and imprinted loci. A knock-in *Asxl3* mouse model harboring a mutation corresponding to one diagnosed in BRS-patient recapitulated the molecular features BRS-patient derived cellular model, including the escape from NMD and Polycomb Repressive Complex 2 (PRC2)-related transcriptomic dysregulation. In contrast, heterozygous *Asxl3* knockout mice and transient knockdown models showed no phenotype, indicating that truncated ASXL3 that may exert dominant-negative effects rather than simple loss of function. This molecular dissection offers new venues for treatment, including allele-specific Antisense Oligonucleotides (ASO), which were used by us in patient-derived cells to downregulate the expression of the mutated allele, and were able to induce partial recovery of the proteomic profile. Taken together, our results support a dominant-negative mechanism for BRS causing truncating mutations, offering a compelling rationale for allele-specific ASO therapeutic strategy.

**One sentence Summary:** Patient-derived neuronal models and mouse models reveal that truncated ASXL3 drive epigenetic dysregulation through a gain-of-function mechanism.

## Introduction

Bainbridge-Ropers syndrome (BRS, OMIM #615485) is a rare, autosomal dominant neurodevelopmental disorder, caused by heterozygous mutations (usually de-novo^1,2^) in the *ASXL3* gene^3^. Patients diagnosed with BRS commonly exhibit global developmental delay, intellectual disability, hypotonia, behavioral abnormalities, and distinct craniofacial dysmorphic features^3–6^. Other symptoms include seizures, breath-holding spells, failure to thrive and more. As sequencing technologies advance, more truncating variants in ASXL3 are identified while expanding the phenotypic spectrum of BRS^2,4^. The underlying pathophysiology and functional impact of *ASXL3* mutations remain poorly characterized, though previous publications suggested Loss of Function (LOF) as a sole mechanism for the disease^7,8^.

*ASXL3* belongs to the additional sex combs-like (ASXL) gene family, alongside *ASXL1* and *ASXL2*, which are critical regulators of gene expression and chromatin remodeling. Members of the ASXL family act as scaffolding proteins that interface with Polycomb Repressive Complex 2 (PRC2) and Trithorax complexes, epigenetic regulators of transcriptional repression and activation, respectively. Both *ASXL1* and *ASXL2* are involved in modulation of histone modification and transcriptional activity; mutations in both genes might result in hematopoietic malignancies, as well as neurodevelopmental disorders (Bohring-Opitz syndrome, OMIM #605039 and Shashi-Pena syndrome, OMIM #617190). Moreover, all ASXL proteins, including ASXL3, can bind to BAP1 and function as a cofactor for the Polycomb Repressive-Deubiquitinase (PR-DUB) complex^9,10^ that removes H2AK119ub1,a ubiquitination mark catalyzed by Polycomb Repressive Complex 1 (PRC1).These processes are essential for maintaining cellular differentiation and developmental programming. Although *ASXL1* and *ASXL2* are well-studied, particularly in hematopoietic malignancies, much less is known about the role of *ASXL3* in normal development and disease. Evidence suggests that *ASXL3* may share functional overlaps with *ASXL1* and *ASXL2*^11^, while maintaining unique roles in neurodevelopment, as it is highly, and almost exclusively, expressed in fetal and adult brain tissue^12,13^.

Understanding the molecular mechanisms underlying rare genetic disorders is critical for the development of effective and tailored therapeutic strategies, particularly in the era of precision medicine and genetic therapy. Advances in gene-targeting technologies, such as antisense oligonucleotides (ASOs)^14^, CRISPR-Cas systems, and AAV based gene augmentation, have opened new possibilities of personalized solutions for disease-causing pathogenic variants^15,16^.

This work aims to explore the molecular effect of truncated *ASXL3* in health and disease, and its relevance in the pathophysiology of Bainbridge-Ropers syndrome. By leveraging both molecular and *in vivo* tools, we seek to clarify how specific *ASXL3* mutations lead to disease phenotypes, shedding light on potential therapeutic approaches for this rare and debilitating syndrome.

## Results

### *ASXL3* c.4826G>A and other BRS-causing mutations escape nonsense mediated decay

*In silico* analysis of reported mutations in *ASXL3* show that BRS patients are usually diagnosed with truncating mutations, resulting in putative Loss of Function (pLOF). These include splicing variants, frameshifts or nonsense mutations, resulting in premature termination codon (PTC). The majority of these mutations are located in exons 11 and 12, the largest exons in the gene, spanning more than 80% of its total length. However, not all truncating mutations result in BRS, as pLOF mutations are also diagnosed in healthy individuals, mainly on the n-terminus, as seen on genetic databases (i.e-gnomad) (**Figure 1A)**.

**Figure 1.**
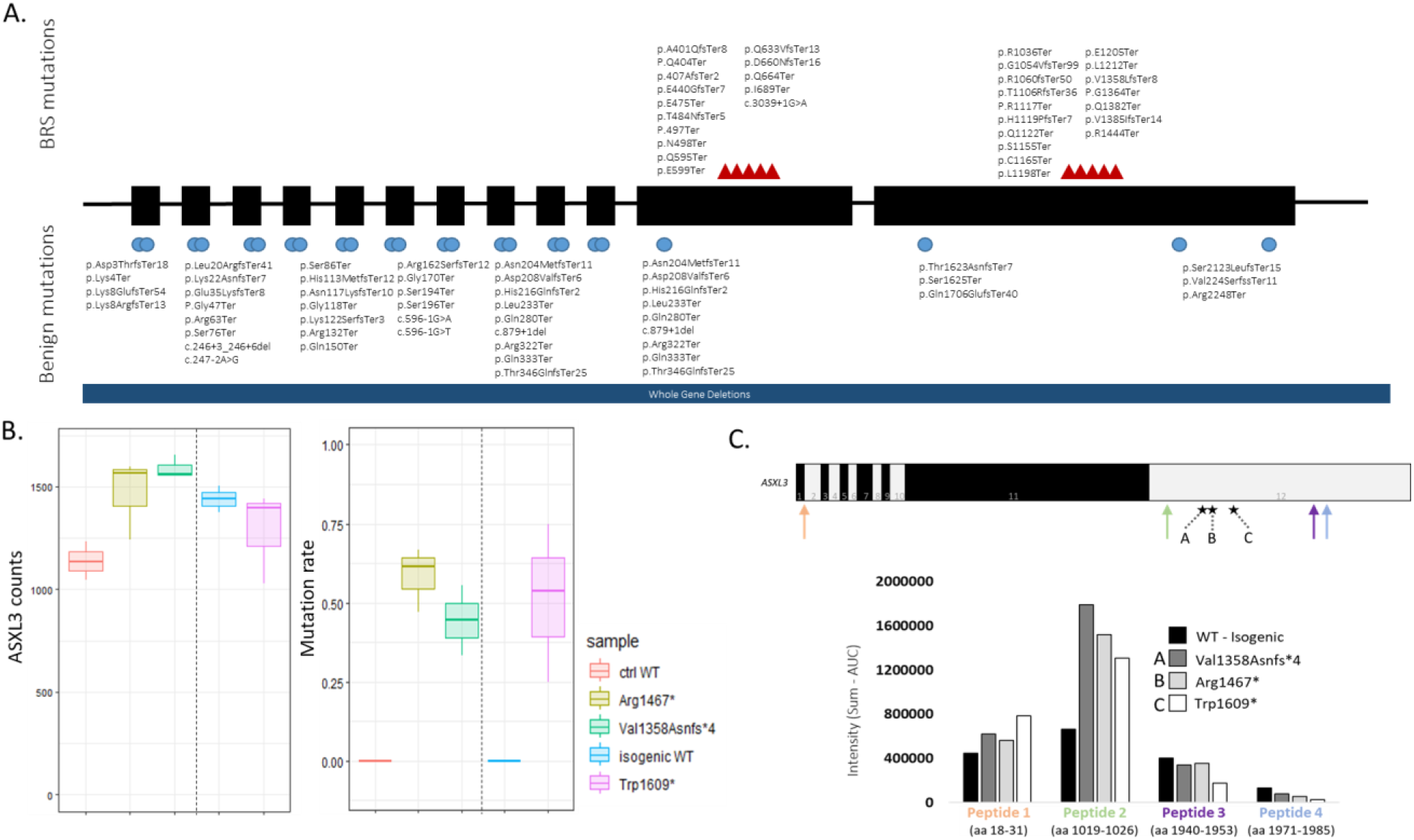
ASXL3 c.4826G>A and other BRS-causing mutations escape NMD. (A) Schematic representation of reported ASXL3 variants. Blue circles indicate benign and likely benign variants; Red triangles indicate known BRS causing mutations. (B) *ASXL3* read counts and calculated mutation rate as seen in RNAseq in patient and control –derived DRG neurons. (C) Illustration of ASXL3 peptides detected by targeted-MS and their intensity in patient and control– derived DRG neurons. Each colored arrow represents the corresponding peptide in the intensity bar graph bellow.

In order to further elucidate ASXL3 pathophysiology in BRS, we aimed to molecularly characterize BRS-causing mutations, utilizing patient-derived cellular models. Our primary focus was a patient with a nonsense mutation in *ASXL3* (NM_030632.2 c.4826G>A, Trp1609*), clinically manifesting as BRS. Fibroblasts from this patient were cultivated and reprogrammed to induced pluripotent stem cells (iPSC). We established an isogenic control (utilizing CRISPR-based Adenine Base Editor), carrying the exact same genetic traits, apart from the pathogenic *ASXL3* variant (referred to as isogenic control). We also received additional patient-derived cell lines (harboring different mutations) from SFARI (Val1357Asnfs*4, Arg1467*, Arg1036*, Asp660Asnfs*16). All cell lines were reprogrammed to iPSC and differentiated to neural progenitor cells (NPCs) and subsequently dorsal root gangilion (DRG) neurons, as *ASXL3* is primarily expressed in neural lineage, and not in fibroblasts (**Figure S1A-B**). DRG neurons of 1609* and its isogenic control, Val1357Asnfs*4, Arg1467* and normal healthy donor (NHD) were subjected to PolyA RNA-seq, and subsequent *ASXL3*-focused analysis. Gene expressions levels were analyzed as total normalized reads count and the mutant allele read counts were calculated relative to all read counts (**Figure 1B)**; in all patient-derived cell lines, results did not suggest nonsense mediated decay (NMD), as the gene expression levels did not significantly differ from the isogenic control and NHD cells. Since all of these mutations are located in the last exon, they are expected to escape NMD^17,18^, aligning with the RNA levels seen in BRS patients’ cells. Similar results were observed in polyA RNA-seq of patient-derived neural progenitor cells (NPCs) (**Figure S1C**), and were validated with Sanger sequencing of the cDNA **(Figure S1D**) and RT-qPCR (**Figure S1E**). Mutant RNA stability was also observed in RT-qPCR and Sanger sequencing of the exon11 truncating variant Asp660Asnfs*16, which was predicted to undergo NMD by its locations, showing no change in *ASXL3* transcript levels (**Figure S1E-F**). Next, we applied targeted Mass Spectrometry to the same samples, showing that ASXL3 peptides in DRG neurons were more abundant in patients compared with controls (isogenic and NHD) (**Figure 1C**), suggesting possible accumulation of the truncated protein, and further excluding possible NMD of the truncated *ASXL3* in these specific patients.

### ASXL3 pathogenic variants alter transcriptome and proteome in patient-derived cellular models

In order to further characterize the molecular phenotype of BRS, we subjected patient derived cells (ASXL3-1609*) and their isogenic control to RNA-seq and mass spectrometry analysis (differential transcriptomics and proteomics). RNA-seq was conducted on DRG neurons, revealing differential expression profile between the lines (**Figure 2A**). GO-enrichment (molecular function) for differentially expressed genes included gene-regulation functions (specifically – DNA-binding transcription activator activity) and E-box binding (**Figure S2A**). Moreover, ChEA^19^ consensus analysis showed enrichment of genes regulated by SUZ12 (FDR 2.8E-06) and EZH2 (FDR 4.3E-11), both components of PRC2. Among the most differential genes, we were able to identify HOX genes (i.e *HOXA2, HOXA3, HOXB4*) and golgin genes (**Figure 2A, S2**C), highlighting ASXL3 importance as an epigenetic regulator in developmental processes. Another significantly enriched gene was the lncRNA *H19* (**Figure S2B-C**). These results were reproducible in additional patient-derived cell lines (ASXL3-Arg1467Ter, ASXL3-Asp66Asnfs*16) (**Figure 2B, S2C)**, validating the contribution of *ASXL3* pathogenic variants to this dysregulation. Proteomic analysis of neural progenitor cells (NPCs) differentiated from ASXL3-1609* and its isogenic control (**Figure 2C**), as well as three additional patient-derived NPCs (ASXL3-1467*, ASXL3- Thr398Profs*10 and ASXL3- Val1358Asnfs*4) revealed an additional layer of differential expression profile, highlighting specific proteins like PDLIM3 and EGFR, which consistently differed between said samples (**Figure 2D**). In unsupervised hierarchical clustering, BRS patients cluster together, validating the contribution of differentially expressed proteins (DEP) to the molecular phenotype (**Figure 2D**). These finding were validated by Western blot analysis (**Figure 2E**), and were consistent in DRG neurons and NPCs (**Figure S2D-F**). Western blot analysis also revealed an increase in BAP1 expression (**Figure S2E**). Interestingly, elevated levels of BAP1 are also observed as a result of truncating mutations in *ASXL1*^20^.

**Figure 2.**
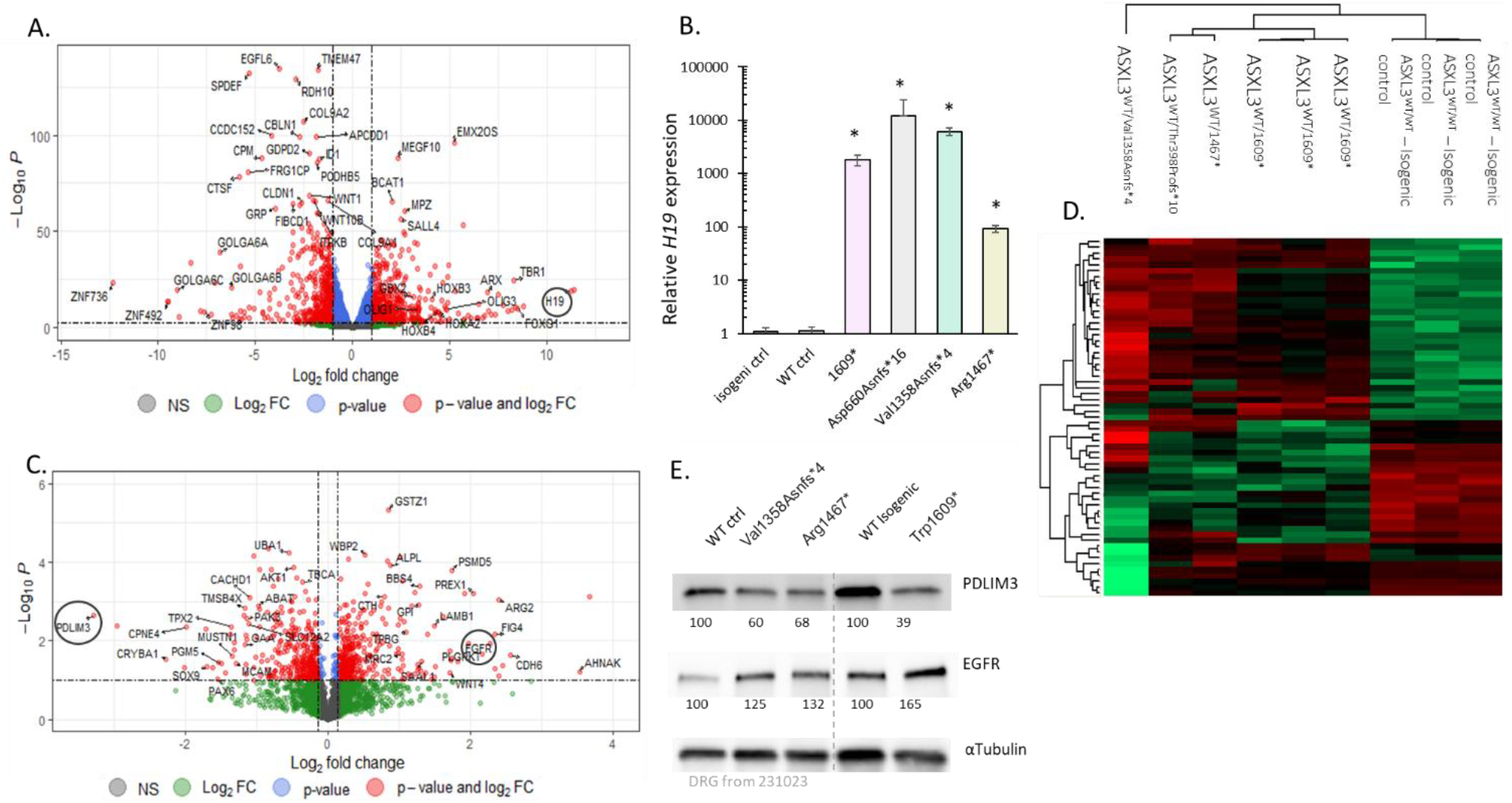
ASXL3 pathogenic variants alter transcriptome and proteome in Trp1609* patient-derived cellular models. (A) Volcano plot of differentially expressed genes as seen by by RNA-seq between Trp1609* patient and control –derived DRG neurons. (B) Transcript level of *H19* using RT-qPCR in patient and control –derived DRG neurons. (normalized to GAPDH, n=3, * p value<0.01). (C) Volcano plot of differentially expressed proteins as seen by mass spectrometry of patient and control -derived NPCs. (D) Heat-map of differentially expressed protein based on mass spectrometry analysis of patient and control –derived DRG neurons. Western blot of PDLIM3 and EGFR differentially expressed proteins in Trp1609* patient and control –derived DRG neurons. α-Tubulin was used as loading control. Band intensity was measured using ImageLab software and normalized to WT control. (E) Western blot of PDLIM3 and EGFR differentially expressed proteins in Trp1609* patient and control –derived DRG neurons. α-Tubulin was used as loading control. Band intensity was measured using ImageLab software and normalized to WT control.

We have also established a bi-allelic isogenic ASXL3-KO by incorporating a premature termination codon predicated to result in NMD (**Figure S3A**), using the original patient-derived iPSC (ASXL3-1609*). Indeed, RT-qPCR showed low residual expression of ASXL3^21^ (**Figure S3B**). ASXL3-/- (also referred to as ASXL3-87*/87*) iPSC failed to properly differentiate to DRG neurons, as demonstrated by RT-qPCR analysis of differentiation markers (i.e *MAP2*, *NEUROD1*, *NSE* etc.) (**Figure S3C**) and morphologically (**Figure S3D**). Moreover, specific RT-qPCR for *H19* showed near-normalized levels of *H19*, comparing to its levels in patent-derived cells lines (**Figure S3E**). Additional analysis by RT-qPCR was conducted using NPCs, showing distinct transcriptomics patterns, including opposite trends in KO compared with ASXL3-1609* (*VCAM1, NCAM2, HOXB5, TXNRD1*), alongside genes which differ only in KO (i.e *COMPT*, *ZNF229*), or genes which show the same trend, but in different magnitudes (i.e *COL9A1, MEG3*) (**Figure S3F**). These findings underline a distinct pathophysiology for the ASXL3-1609* mutant allele, which differs from simple loss of function.

### Altered chromatin accessibility and epigenetic landscape in an ASXL3-1609* cellular model

H2AK119ub levels were assessed in patient-derived NPCs and their isogenic controls (WT and -/-) by Western blot. The patient cell line exhibited slightly reduced H2AK119Ub compared to WT, while the KO isogenic line showed increased levels of K119Ub1 relative to WT (**Figure 3A**). These finding are consistent with *ASXL3* function as a part of PR-DUB^9^, and support the notion that ASXL3-1609* acts as a neomorph, with distinct function from simple loss of function.

**Figure 3.**
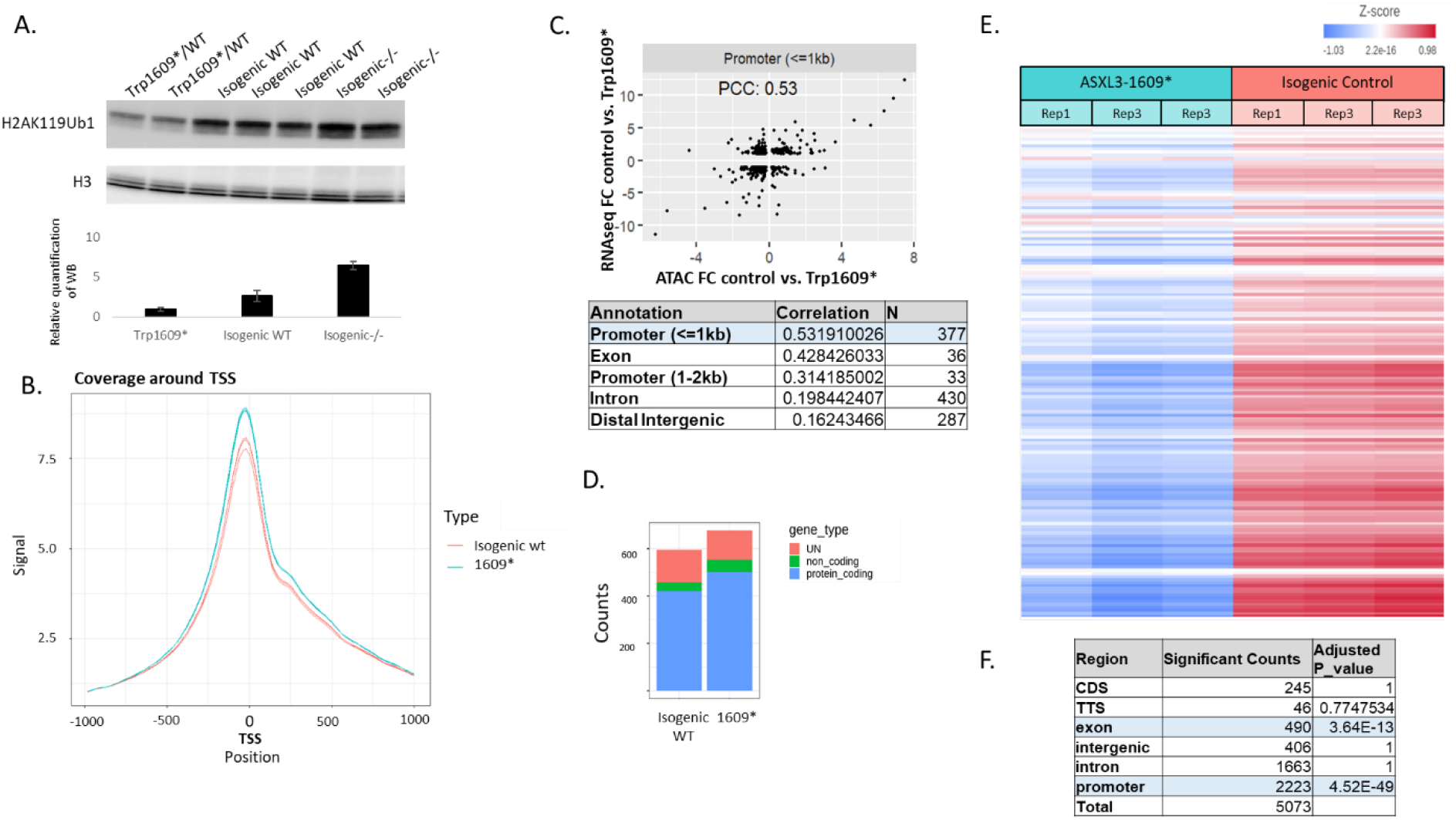
Altered chromatin structure and epigenetic signature in cellular models. (A) Western blot of H2AK119ub levels in patient-derived DRG neurons, isogenic control and bi-allelic KO. Anti-H3 was used as loading control. (B) Reads coverage around the transcription start site (TSS) as identified by ATAC-seq on patient-derived and isogenic control DRG neurons. Signal intensity is calculated using recoup package (Bioconductor). (C) Correlation between differentially expressed genes seen in RNA-seq and promotor accessibility in ATAC-seq. (D) Read counts in the regions of open chromatin as identified by ATAC-seq analysis on patient-derived and isogenic control DRG neurons. (E) Differentially methylated regions heatmap for patient-derived DRG neurons, comparing to their isogenic control. (F) Distribution of differentially methylated probes, P value was calculated in Fisher’s exact test.

Next, chromatin accessibility was measured by ATAC-seq conducted on patient-derived and isogenic control DRG neurons, showing that the chromatin in ASXL3-1609* cells is more accessible (“open”) (**Figure 3B**). ATAC-seq results were highly reproducible, and revealed distinct chromatin accessibility patterns between the experimental groups **(Figure 3B**). Accessible regions were enriched mainly in promotors (**Figure S4A**), and the peaks were correlated with RNA-seq data (PCC=0.53) (**Figure 3C**) and with overall higher expression rates, as shown by RNA-seq analysis (**Figure 3**D). GO analysis (molecular function and biological processes) for differentially enriched regions show enrichment in genes related to signaling (G-protein coupled receptor) and synaptic activity (**Figure S4B-C**). Pathways analysis (KEGG) showed specific enrichment of axon guidance, but also cell adhesion (**Figure S4D**). Consistent with high chromatin accessibility, methylation analysis of over 935,000 probes (EpicV2) shows a dramatic hypomethylation of patient-derived DRG neurons, compared with their isogenic control (**Figure 3E**). Differentially methylated probes were located mainly on exonic regions and promoters (**Figure 3F**), and enriched for transcription regulation and gated channel activity (GO annotations, molecular function), as well as “neuron differentiation”, “cell projection organization”, “cell motility” and more (GO annotations, Biological processes) (**Figure S5A**). A significant enrichment was also seen for probes located on the X Chromosome, suggesting aberrant X inactivation (Odds ratio = 11.93, Fisher’s Exact Test, P.Value<0.00001) **(Figure S5B**). An additional noteworthy observation was seen upon analyzing the differentially methylated regions based on transcript type. These regions demonstrated significant enrichment in lncRNAs (Fisher’s Exact Test, p-value = 4.55 × 10⁻¹⁶) **(Figure S5C**). Further analysis of significantly hypomethylated regions revealed significant enrichment for imprinted genes (Fisher’s Exact Test, P.Value = 0.01). This notion was strengthened by detection of bi-allelic expression from imprinted genes (**Figure S5D)**, utilizing data from RNA-seq, suggesting disturbance in methylation and imprinting.

Analysis of additional histone modifications in DRG neurons by ModSpec revealed no significant changes in the vast majority of assessed histone marks (H2AK119Ub1 not included), apart from H3K79me1, H3K9ac, H4K20ac – all marking active transcription and an open chromatin (**Figure S5E**).

### Allele-selective reduction of mutant *ASXL3* as a therapeutic approach

As our work demonstrated that some of the BRS-causing pathogenic variants does not result in NMD, we decided to harness allele-selective, RNAse-H1 recruiting ASOs, to reduce the expression of the mutant allele, in patient-derived cells harboring the c.4836G>A mutation (ASXL3-1609*). To that end, we designed a library of ASOs specific to the mutant allele, either by directly targeting the mutation or by leveraging linked SNPs. We conducted a preliminary screen using a luciferase-based plasmids, expressing either the WT or mutant version of the gene (**Figure 4A**). Efficient and selective ASOs were also subjected to toxicity prediction assay^22,23^, in order to eliminate potentially pro-inflammatory ASOs (**Figure S6A**). Patient-derived DRG-neurons were then treated gymnotically with leading, non-toxic ASO candidates, showing efficient and selective reduction of the mutant allele (**Figure 4B, S6B**). Subsequent analysis of differentially expressed proteins, including PDLIM3, showed rescue of the phenotype following a 5-day treatment with allele-selective ASO (**Figure 4C**).

**Figure 4.**
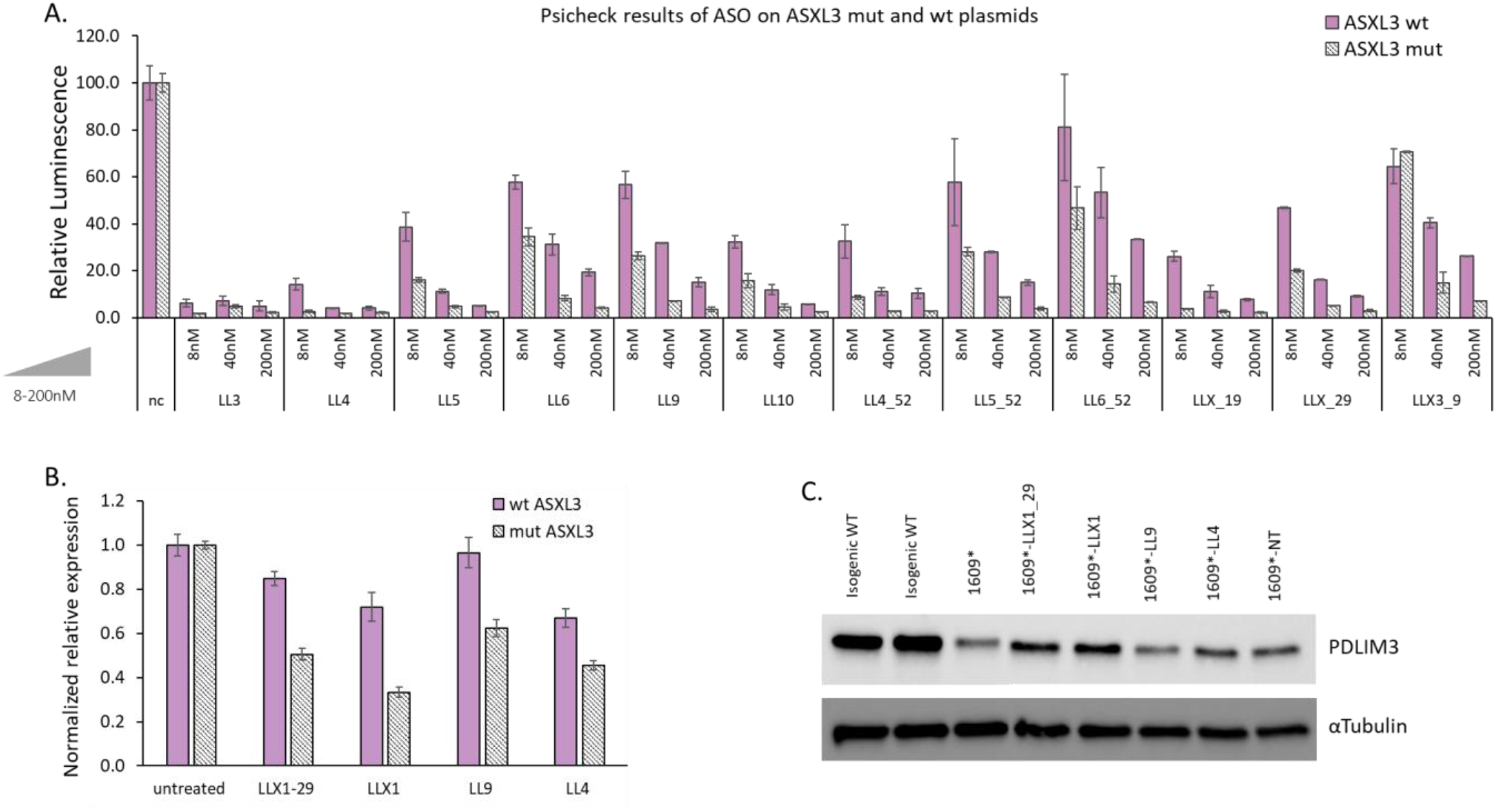
Allele-selective reduction of mutated ASXL3 as a therapeutic approach. (A) ASO-Screen using luciferase-based plasmids. Luciferase levels of WT and MUT plasmids were measured following ASO treatment with increasing concentration of ASO (8nM- 200nM ASO, co-transfected with MUT or WT psi-CHECK plasmids). n=3 (B) *ASXL3* WT and mutant transcripts quantification using rtPCR in patient-derived DRG neurons treated gymnastically with selected ASO for 72 hours. (C) Western blot of PDLIM3 in isogenic control and patient –derived DRG neurons following ASO treatment for 5 days. α-Tubulin was used as loading control.

### Asxl3-1556* BRS model mouse recapitulates the human BRS molecular phenotype

In order to further elucidate the molecular mechanism underlying BRS, and validate ASOs as a therapeutic approach, we established a mouse model. This new strain carries a mutation in murine *Asxl3*, which corresponds to the ASXL3-1609* patient mutation (referred to as Asxl3-1556*) (**Figure S7A**), established using CRISPR/Cas9 injection to blastocyst (C57BL/6JOlaHsd background). Heterozygous mice were viable and fertile, showing no gross anatomical anomalies (**Figure S7B**). Importantly, relative expression of *Asxl3* (total, WT and mutant alleles) in cortices harvested from p21 heterozygous mice showed that the mutant allele did not undergo NMD (**Figure 5A**), thus recapitulating the molecular mechanism of the human corresponding mutation. Growth analysis showed that Asxl3-1556*heterozygotes were slightly smaller than WT littermates-an effect which was seen in both male and female, but more profoundly in males (**Figure S7C**). Homozygous mice were detected in utero at day e12.5 and as p2 neonates at the expected mendelian ratios, but most did not survive to weaning (resulting in 2.1% homozygous mice at day 21) (**Figure S7D**). This outcoe could be partially attributed to maternal care malfunctions, as reproduction follow-up revealed that female heterozygotes gave fewer litters, significantly smaller, and many did not survive until weaning; more HT females lost the entire litter (27.3%), comparing to WT littermates (6.7%) (**Figure S7E**). Next, comprehensive behavioral and electrophysiological characterization of Asxl3-1556* heterozygote mice was conducted and compared to WT controls (littermates) across two time points (ages 5 and 9 weeks). Twenty mice, heterozygote and WT, were divided into four groups in accordance with gender and genotype (n=5 per group). All mice were subjected to a battery of standardized assays assessing motor function (Catwalk gait analysis, Rotarod, Open Field Test), sensory response (Von Frey, Hot Plate, Cold Plate), cognitive performance (Morris Water Maze, Elevated Plus Maze), and electrophysiological parameters (tcMEP, CMAP, EMG, tcSEP, VEP). Behavioral assessments were performed at both time points, allowing longitudinal comparison within and between groups. The electrophysiological recordings were performed only on 9-week old mice, and enabled detailed analysis of central and peripheral nerve function, including motor, sensory, and visual pathways. The same groups were later euthanized and subjected to exploratory anatomical dissection and histopathology of hearts and brains. All data sets were analyzed using appropriate statistical methods, showing no differences between the genotypes in most of the tests (**Figure S7B**). Minor changes were seen in motor tests, as the heterozygotes had a slight better overall performance. Next, we sought to further characterize these mice, using RNA-seq conducted on cortices of 21-day old female mice. Differentially expressed genes were then subjected to ChIP-X Enrichment Analysis (ChEA)^19,24^, showing enrichment of genes that are regulated by *Jarid2, Suz12,Rnf2, Ezh2* and *Eed* – all components of the PRC2 complex (**Figure 5B-C**). These results indicate that the aberrant transcriptomics in Asxl3-1556* heterozygous mice might stem from PRC2 altered activity, possibly due to Asxl3 malfunction.

**Figure 5.**
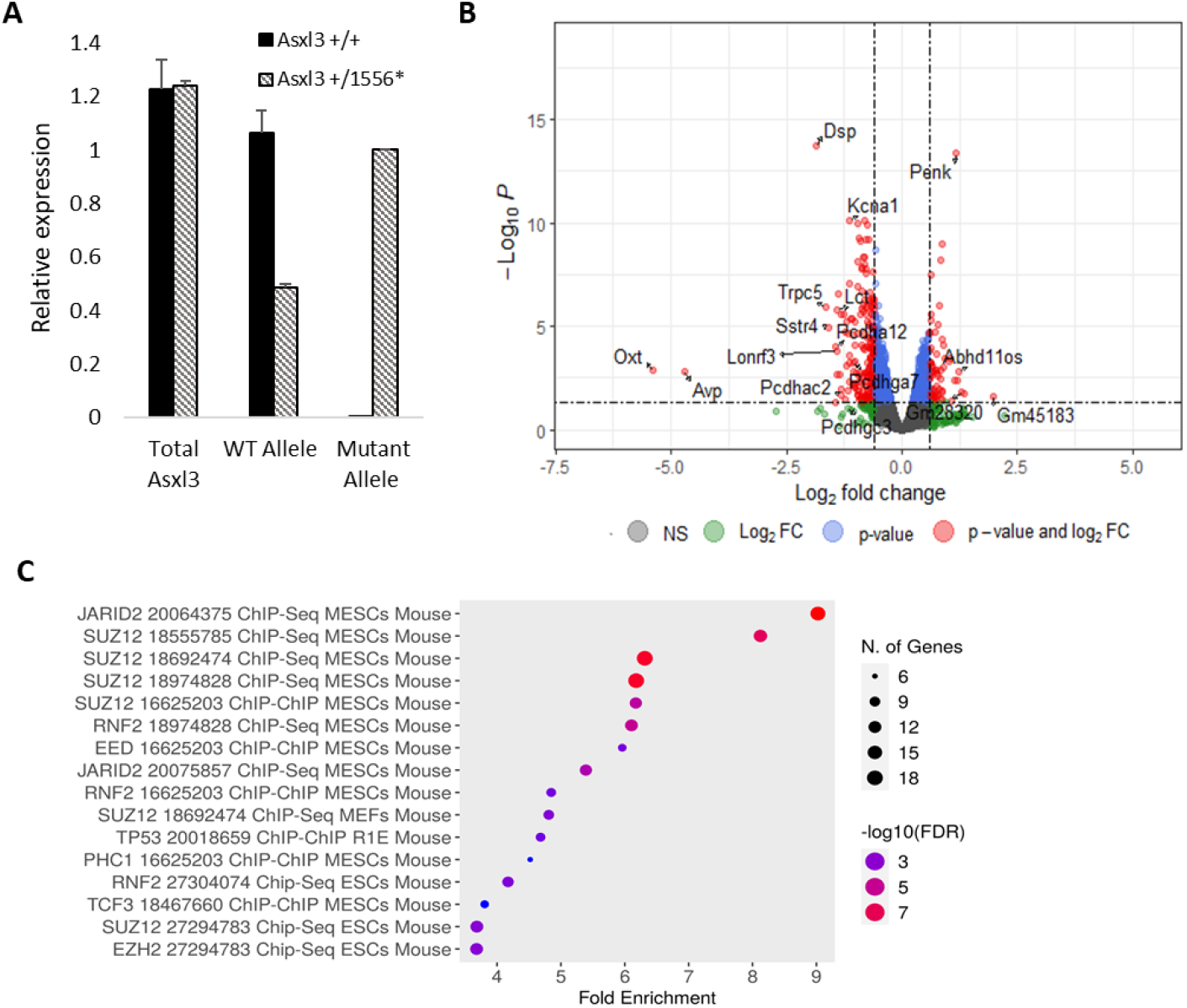
Asxl3-1556* mouse model recapitulates molecular traits of the human model. (A) Asxl3 transcript analysis in cortices harvested from p21 female heterozygote Asxl3-1556* mice shows that the mutated allele escapes NMD (n=3). (B) Volcano plot of differentially expressed genes in RNA-seq of cortices in adult female Asxl3-1556* heterozygote comparing to WT littermates (n=3). (C) ChEA enrichment for differentially expressed genes (padj< 0.05, log2(FC) > 1) between Asxl3-1556* and WT littermates.

### Heterozygous Asxl3 knockout mice show no neurological phenotype

Next, we sought to evaluate the effect of Asxl3 deletion on the development and behavior of mice. To that end, we have established an additional mouse model, harboring a deletion of Asxl3-Exon11, predicted to result in PTC and subsequent NMD. Asxl3-KO was validated by mass spectrometry for brains of P0 homozygotes, compared with WT littermates, showing no Asxl3 peptides identified in homozygotes’ brains (**Figure 6A**). Heterozygote carriers of the deletion (referred to as Ht Asxl3-Del11 or +/-) were viable and fertile, and showed no significant changes in overall appearance and behavior in cage-side observations. Comparative transcription analysis using RNA-seq was performed on heterozygote and WT littermates (n=3), showing no significant differences (**Figure S8A**). This contrasts with differentially expressed genes (DEG) observed when comparing Asxl3-1556* heterozygotes and WTs. Electrophysiological recordings (VEP, ENG and BERA) performed on 9-week-old male mice (n=5) showed no significant difference between heterozygotes and WT littermates (**Figure S8B**), suggesting normal function of the central and peripheral nerve system. Homozygous mice were not seen at the expected mendelian ratios, with almost no homozygous pups born (**Figure 6B)**. Surviving homozygous mice were significantly smaller and less developed than wildtypes and heterozygous littermates (**Figure 6C-D**). In terms of survival rates, homozygous Asxl3-Del11 showed a dramatic and significant decrease in lifespan when measured from birth, showing median survival rate of 24 hours (**Figure 6E**). When measured from weaning, homozygous mice showed overall similar survival to heterozygous and WT littermates (**Figure 6F**), suggesting pre-weaning lethality.

**Figure 6.**
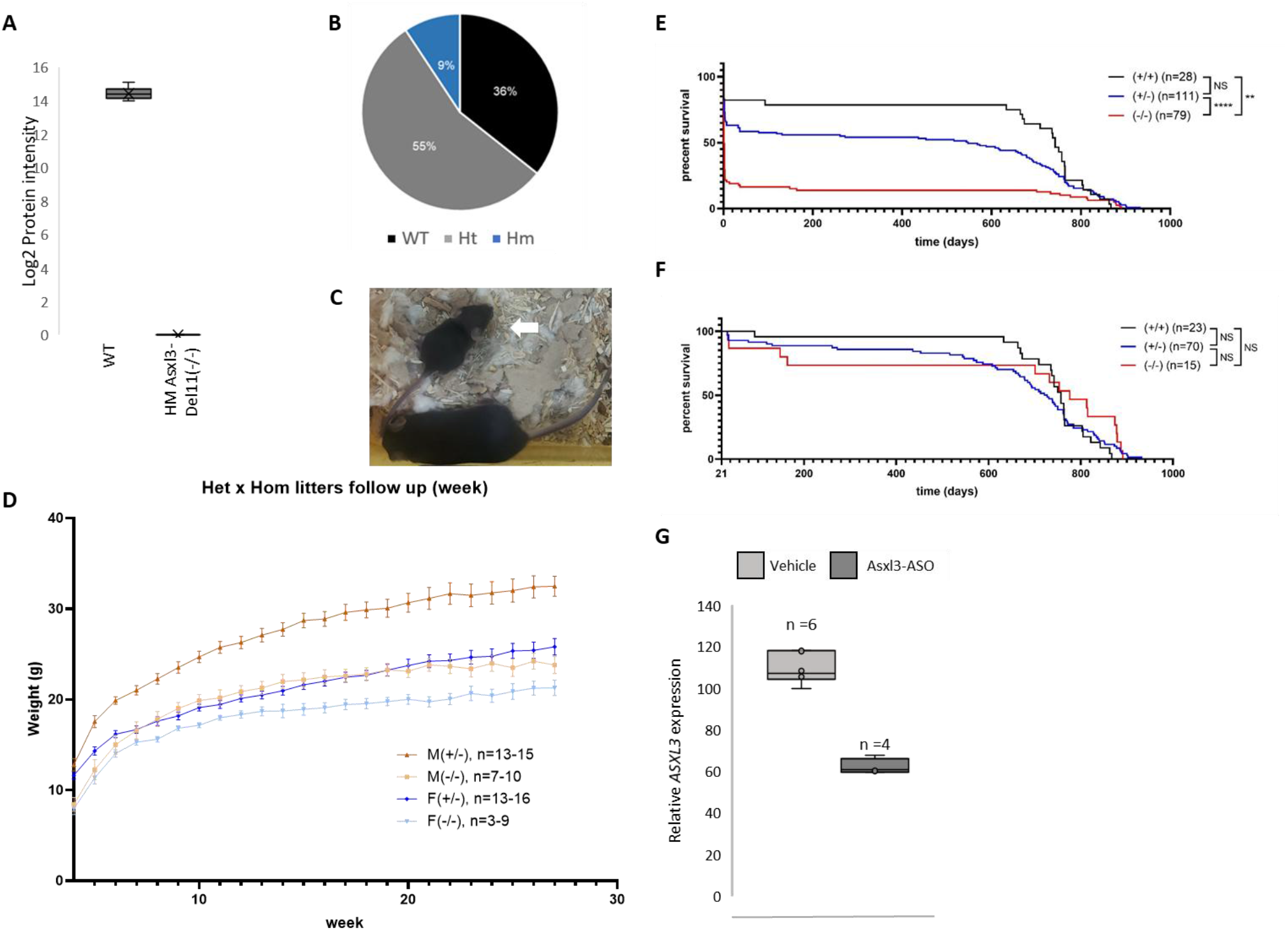
Heterozygote knockdown and genetic knockout of mAsxl3 is tolerated in mice. (A) Mass Spectrometry analysis for Asxl3 protein abundance in P0 brain tissue harvested from homozygote Asxl3-Del11(-/-) and WT littermates validates Asxl3 knockout. (B) Pie chart of pups distribution for Asxl3-Del11 strain, in HTXHT breeding (n=129, χ^2^= 6.66E-05). (C) Representative picture of significantly smaller homozygote Asxl3-Dell1 pup (-/-, marked by white arrow) next to a heterozygote littermate (age = p17). (D) Growth curves for homozygotes Asxl3-Del11 (-/-) and heterozygote littermates (+/-) during weeks 4-27. For all time points, p value for males <0.001, p value for females is <0.05 (E) Survival curve of Asxl3-Del11 mice (+/- and -/-, male and female) from birth. (F) Survival curves for Asxl3-Del11 (+/- and -/-, male and female) from day 21 (weaning), **<0.005, ***<0.0001) (G) Asxl3 transcript levels 10 days after injection of PBS (10ul) or mAsxl3 targeting RNase-H1 recruiting ASO (6mg/kg, 10ul) in WT C57BL/6J, 8-weeks old mice (female) (** p-value< 0.01).

In addition to the genetic deletion of Asxl3, which showed no significant phenotypes in heterozygotes, we used ASOs targeting Asxl3 to transiently reduce its expression in 8 week-old WT mice (C57BL/6J). Different ASOs, designed to target the human and murine sequences of *ASXL3* were screened using 3T3 cells, and lead candidates were further tested for dose-response correlation (**Figure S8C**). The most efficient ASO (mhASO8) was selected and administered via intracerebroventricular (ICV) injection (6mg/kg). Mice were monitored for 10 days post-injection, during which all mice gained weight and showed no behavioral abnormality in cage-side observations. RT-qPCR analysis revealed 40% decrease in *Asxl3* expression, which did not result in any observable phenotype (**Figure 6G**). Subsequent PolyA RNA-seq analysis revealed 175 differentially expressed genes, including more than 15 protocadherins (both α and γ clusters) (**Figure S8D**), which were also highlighted as differential between Asxl3-1556* heterozygotes and littermates (**Figure 5B**).

The same ASO (mhASO8) was also administered via ICV injection to male heterozygous Asxl3-1556* mice (n=6 per group, 6mg/kg). The experimental period lasted 28 days following ASO administration. Throughout the study, no procedure-related phenotypic changes were observed. After euthanization, brains were harvested, and subsequent transcriptomic analysis revealed a decrease in both WT and mutated Asxl3 alleles (20-30%) (**Figure S8E**). In addition, genes previously identified as responsive to Asxl3 reduction via ASO treatment in WT mice (**Figure S8D**) showed consistent upregulation, as confirmed by RNA-seq analysis in heterozygous mice (**Figure S8F**).

## Discussion

BRS is a rare neurodevelopmental disorder, which stems from pathogenic variants in ASXL3, an epigenetic regulator. ASXL3 belongs to the ASXL family of proteins, which interact with chromatin-modifying complexes^11^, including PRC1, PRC2, PR-DUB and Trithorax Group. During development, ASXL3 is highly expressed in fetal and adult brain tissue, where it presumably holds a crucial role in regulating gene expression programs essential for proper neuronal differentiation and function^10,12,25^. Being a rare disorder, the pathophysiology of BRS remains poorly characterized; in this work, we set out to describe the molecular phenotype of ASXL3 pathological variants, in patient-derived cell lines and mouse models. So far, BRS was considered as stemming exclusively from ASXL3 haploinsufficiency^7^, mainly due to the fact that all BRS causing mutations that were reported up to date were truncating variants, including nonsense mutations and frameshifts.

Studies have shown that truncating mutations in other members of the ASXL family, *ASXL1* and *ASXL2,* lead to the loss of proper protein function in addition to dominant-negative effects^26–29^, interfering with the activity of remaining wild-type protein. *ASXL1* truncation disrupts crosstalk with PRC2 and Trithorax complexes^30^, leading to widespread alterations in gene activation and repression. We speculated that a similar mechanism may underlie *ASXL3*-mediated disease, where truncating mutations prevent normal chromatin remodeling and gene regulation, thereby impairing neuronal development and homeostasis.

As mentioned, many of the BRS causing mutations result in PTCs. Such events can trigger NMD, a surveillance pathway that eliminates transcripts with PTCs, hence preventing the accumulation of truncated proteins, resulting in reduced expression of the gene. In order to trigger the NMD circuit, the PTC must be located upstream of exon-junction complexes (EJCs), which can be usually found within 20-24 nts upstream of exon-exon splicing junctions. Since PTCs in the last exon lack downstream exon-exon junctions, there are no associated EJCs to facilitate the recruitment of the NMD machinery, resulting in escape from NMD^18,31^. This is an important notion to consider when examining BRS-causing mutations, as many of these are found in the last exons (11-12). In the work presented hereby, we molecularly examined different BRS-causing mutations, and concluded that they most likely result in an escape from NMD and production of a truncated protein. Such truncation can result in a protein lacking specific domains, like the c-terminal DNA binding domain. These results suggest that BRS does not stem solely from haploinsufficiency, and could be mediated by additional mechanisms, including dominant-negative, or neomorph expression^17,32^. Multiple genetic mechanisms are usually correlated with distinct phenotypes; However, while BRS clinical phenotype is variable, there is no known phenotype-genotype correlation^4,5^. Additional work will be required to determine whether such correlation exists, or whether these different mechanisms somehow converge into a shared pathology. In the scope of this work, we chose to focus on variants that escape NMD^17,31^, and specifically ASXL3-1609*, a variant diagnosed in a BRS patient presenting a global developmental delay, hypotonia and language impairment.

ASXL3 is a member of the PR-DUB complex, mediating the de-ubiquitination of histone H2A lysine 119 (H2AK119Ub1)^7,9^. Interestingly, our results showed a slight decrease in H2AK119ub1 levels in patient-derived NPCs, distinct from accumulation of H2AK119Ub1 in ASXL3-KO NPCs, compared with the isogenic control.

*ASXL3* mutations studied in this work significantly altered the transcriptome, epigenetic landscape and chromatin status, and proteome both in patient-derived cells and in the mouse model. ATAC-seq analysis for patient-derived DRG neurons and their isogenic controls showed increased chromatin accessibility in patient samples. While such an effect can be attributed to aberrant differentiation and retainment of plutipotency, major pluripotency regulators (including *KLF2, KLF17, DPPA3, DPPA5, NANOG*, and *POU5F1*^33^) were not found in the differentially accessible regions, and GO analysis showed enrichment for signaling pathways and adhesion molecules. These notions were further supported by detection of hypomethylation in patient-derived DRG neurons, with differentially methylated regions enriched for transcription regulation, DNA binding and nervous system development.

These findings align with previous publications describing ASXL3 function as an adaptor protein, that can form a bridge between chromatin modifiers and chromatin binding proteins, like BRD4^34,35^. The disruption of this function by truncated ASXL3 proteins likely contributes to the altered chromatin accessibility and methylation patterns, as shown in this study. Transcriptional-related changes were also seen in our BRS mouse model, showing enrichment of differentially expressed genes regulated by components of the PRC2 complex. Previous publications showed that both ASXL1^36^ and ASXL2^37^ can directly bind EZH2 and thus to mediate PRC2 binding to its targets; this binding is mediated by N-terminal domains^36,37^, which are well conserved between the ASXL proteins, including ASXL3. Another piece of evidence suggesting a potential link between ASXL3 and PRC2 is the dramatic upregulation of HOXA genes observed in patient-derived cells, as these genes are typically repressed by PRC2^38,39^.

Antisense Oligonucleotides (ASOs) have emerged as a therapeutic promise, particularly for genetic neurological disorders.

As we hypothesized that ASXL3 truncating mutations can result in a dominant negative effect, we harnessed RNAse-H1 recruiting ASOs, as a therapeutic approach for BRS. We designed allele-selective ASOs, specifically targeting the mutant allele, while preserving the WT allele expression in patient-derived neuronal cells. This approach resulted in the rescue of a specific molecular phenotype: the normalization of differentially expressed proteins like EGFR and PDLIM3. In order to further explore ASXL3 haplosufficiency, we established a new m*Asxl3*-KO model-showing no phenotype in heterozygotes. We have also downregulated Asxl3 expression by an ICV injection of bi-allelic Asxl3-targeting ASO, resulting in approximately 40% reduction in its expression-with no visible consequences, tested by transcriptomic analysis and cage-side observations. These findings also suggest partial functional redundancy with *ASXL1* and *ASXL2*, both expressed in the CNS. It is plausible that *ASXL1* and/or *ASXL2* can compensate^40,41^ for *ASXL3* reduced expression.

Taken together, we speculate that allele-selective reduction might be clinically beneficial for BRS patients. Translation of these findings to clinical application will involve design and manufacturing of individualized ASOs for different mutations, as done today for other indications as personalized treatments^42–45^.

## Materials and Methods

### Study Design

This study was designed to define the molecular mechanism by which disease associated truncating variants in *ASXL3* cause Bainbridge-Ropers syndrome, and to evaluate whether this mechanism could support an allele-selective therapeutic approach. We combined patient-derived cellular models, isogenic controls, and complementary mouse models to define mutant *ASXL3* function. Patients’ fibroblasts carrying pathogenic *ASXL3* variants were reprogrammed into iPSCs and differentiated into neural progenitor cells and neurons, enabling transcriptomic, proteomic, chromatin accessibility, DNA methylation, and histone-mark analyses in disease-relevant cellular contexts. To assess causality, we compared these models with CRISPR-corrected isogenic controls, bi-allelic ASXL3 knockout cells, an Asxl3 knock-in mouse model carrying a patient corresponding truncating mutation, and heterozygous Asxl3 loss-of-function mice. Finally, allele-selective gapmer ASOs were designed and tested in patient derived neurons to determine whether selective reduction of the mutant ASXL3 transcript could reverse disease-associated molecular phenotypes.

### Reprogramming of fibroblasts to iPSCs

Patient fibroblasts were derived from a skin punch biopsy (IRB 6158-19-SMC), and reprogrammed to iPSC utilizing StemRNA 3rd Gen Reprogramming Kit (Stemgent) as previously described^45^. Normal human donor iPSCs (KYOU-DXR0109B) were obtained from ATCC (ctrl WT).

### Differentiation of iPSCs to DRG neurons and ASO treatment

iPSCs were differentiated to DRG neurons based on procedure published by Ronald S. Goldstein and modified as previously described^45^. Briefly, iPSCs were seeded in 24 hollow agar molds at a density of 400,000 cells per mold. Cells were grown in GMEM based medium supplemented with 10% KO replacement serum, 1% L-Glutamine,1% pyruvate, 1% non-essential amio acids, 0.1µM β-merchaptoethanol and 1% PCA (Gibco). 20uM Dorsomorphin (Tocris), 10uM SB431542 (Miltenyi Biotec) and 10uM Rock Inhibitor (Enzo Life Sciences) were added during the first 4 days. 14 days following the initial seeding EBs were formed and seeded on Poly-D-lysine/ Laminin (sigma) 24 plates. The neurons were further differentiated for 7 days in DRG medium containing DMEM/F12,2% B27 (Gibco) and 10ng/µl of NGF, NT3, GDNF and BDNF (Alomone). For ASO screens, gymnotic uptake was applied, followed by RNA extraction at day 7.

### Differentiation of iPSC to Cortical neurons

iPSCs were differentiated to cortical neurons based on procedure published by Alessandro^46^. Briefly, 500.000 iPSCs were seeded on Geltrex coated 6 well plates in Nutristem medium supplemented with 10 uM Rock inhibitor. The following day, medium was replaced to fresh Nutristem. 48 hours post seeding cortical induction was started by changing to N2B27 medium composed of 50% Neurobasal, 50% DMEM/F12, 1% Glutamax, 1% non-essential amio acids, 1% Pen/Strep (gibco) and supplemented with 10 uM SB and 500 nM LDN. The media was replaced every day with fresh SB and LDN for 8 days. On day 10 following seeding cell clamps were dissociated using dispase (gibco) and seeded at 1:4 dilution in Poly-L-Ornhitine / Laminin (sigma) 6 well plate with N2B27 medium. On day 20 cell were dissociated using accutase (gibco) and replated on 24 well Poly-L-Ornhitine / Laminin coated plated in N2B27 medium. 180,000 cells per well. For ASO treatment, gymnotic uptake was applied 3 days post seeding, and followed by RNA and protein extraction at day 27.

### qRT-PCR

Cell RNA was extracted using ReliaPrep(TM) RNA Cell Miniprep System (Promega), according to the manufacturer’s instructions. 1000 ng of isolated RNA of each sample was reverse transcribed to cDNA by using High Capacity cDNA RT Synthesis Kit (Applied biosystems). Quantitative real-time PCR was performed on BioRad CFX96 system in technical triplicate per sample by adding 5ul cDNA (5 ng/uL) and primer pairs (listed in **Supplementary Table 1**) to SYBR Green Master Mix (Applied Biosystems). *GAPDH* was used as a housekeeping gene and relative quantification (RQ) values (RQmin/RQmax) were determined using CFX Maestro system. qRT-PCR for differentiation markers and ASXL3 levels were conducted for four independent differentiation experiments, each one includes 3 biological repeats, all quantified in technical triplicates.

### ASO design

ASOs used in this study were synthesized by Ella Biotech (PS-MOE ASOs) and include PS backbone modifications and flanking 2-MOE residues in a gapmer structure. ASOs were diluted to 100 µM stock in DDW, and used as indicated for each experiment. ASOs sequences and modifications are listed in **Supplementary Table 2**.

### psiCHECK

30bp fragment of ASXL3, flanking the mutation site was sub-cloned to psiCHECK™-2 vector (Promega) by PCR and fused to synthetic Renilla luciferase reporter. This vector contains a secondary firefly reporter expression cassette to normalize the relative plasmid quantity.

ASXL3 wild-type fragment: CAAGTAACCGGATTTGCTGGAATGATGATGGGATGAG ASXL3 mutant c.4826G>A fragment: CAAGTAACCGGATTTGCTAGAATGATGATGGGATGAG Luminescence assay was performed as previously described^45^. Briefly, 293T cells were seeded at density of 7 ×10^4^ per well in a white 96 culture plate. Each well was transfected using lipofectamine3000 (ThermoFisher) with 150ng wild-type or mutant ASXL3 pSI-CHECK plasmid and control or ASXL3 ASO in the indicated concentrations. Luminescence was measured 2 days following transfection using Dual-Glo Luciferase assay system (Promega). ’Relative luminescence’ was calculated as the ration of renilla to firefly luciferase in each well. Normalized read was calculated as the ration of ’relative luminescence’ in treated vs. control (untreated) wells.

### Correction of patient derived iPSC to Isogenic WT cells

Isogenic control iPSC for c.4826G>A, Trp1609* patient, were generated using adenine base editing (ABE). Guide sequence duplex containing BbsI overhangs was inserted into pKLV-U6gRNA(BbsI)-PGKpuro2ABFP vector (Addgene #50946) based on Zhang lab procedure. ABE8e plasmid (Addgene # 138489) containing ecTadA (8e)-nSpCas9 was used to express adenine editor. Following transfection of patient’s iPSC using Lipofectamine Stem reagent (gibco), puromycin selection was applied, and single colonies were Sanger sequenced to detect pure isogenic wild-type iPSCs.

Guide duplex cloned to pKLV

Sense- CACC<u>G</u>TTGCTaGAATGATGATGGGAGT

Antisense- TAAAACTCCCATCATCATTCtAGCAA<u>C</u>

### PolyA RNAseq

RNA was extracted from fibroblasts, DRG neurons (day 7) or NPCs as described previously. mRNA was isolated using NEBNext® Poly(A) mRNA Magnetic Isolation Module (NEB) followed by NEBNext® Ultra™ II Directional RNA Library Prep Kit for Illumina® (NEB) for library preparation. Libraries were sequenced on an Illumina NovaSeq 6000 sequencer (Illumina, United States) to generate paired-end (100 bp) reads for each sample. RNA-seq read quality was evaluated using FastQC and adapter sequences were removed with Trim Galore. Reads were aligned to the human genome (GRCh38) using STAR followed by PCR duplicates removal by the unique molecular identifiers (UMIs). Alignment quality was evaluated using Picard Tools. HTseq-count was used to count the number of reads for each gene after alignment. Differential expression analysis was performed with ‘DEseq2’ package in R. GO term enrichment analysis and volcano plot was performed using the ‘clusterProfiler’ and ‘EnhancedVolcano’ packages in R, respectively.

### Western blot

iPSC-derived neurons were lysed in RIPA buffer (Sigma-Aldrich) supplemented with 4% cOmplete protease inhibitor cocktail (PI) (Roche) for 15 min. Cell debris was pelleted at 12,000g at for 15 min, and the supernatant protein content was analyzed. Protein concentration was determined using the QPRO-BCA Kit standard (CYANAGEN).10µg of protein was eluted, mixed with 4X Laemlli sample buffer (BioRad) and heated to 95°C. Samples were loaded and separated on 4-20% polyacrylamide gels in Tris/Glycine/SDSr unning buffer (BioRad). Proteins were transferred onto a nitrocellulose membrane (BioRad) and blocking was performed in 5% skimmed milk in TBS-T. Primary GNAO1 (cst-3975, cell signaling) antibody was diluted 1:1000 in blocking buffer and incubated overnight at 4°C. GAPDH (5174, cell signaling) 1:2000 was used as loading control. After three washes with TBST, the membrane was incubated 1 hour with Rabbit HRP-conjugated secondary antibody (ab6721, abcam). Proteins were visualized using the WESTAR ONE ECL PLUS (CYANAGEN) and Bio-rad imager.

### Histone extraction

Protocol was adapted from abcam protocols for acid histone extraction. In brief, DRG day 7 neurons were harvested and washed with cold PBS containing 5 mM sodium butyrate. Cells were re-suspended with Triton Extraction Buffer (TEB: PBS containing 0.5% Triton X 100, 2 mM phenylmethylsulfonyl fluoride (PMSF), 0.02% NaN3 and 5 mM sodium butyrate) at a cell density of 10^7^ cells per ml, and incubates for 10 min on ice. Cells were centrifuged at 650 x g for 10 min at 4°C and supernatant was discarded. Nuclei pellet was washed twice with TEB, and centrifuged as before. Washed pellet was re-suspended in 0.2 N HCl over night at 4°C (acid extraction). Pellet was centrifuged as before and histone containing supernatant was neutralized using 2M NaOH at 1/10 ratio.

### ATAC-seq

Briefly, DRG day 7 neurons were washed twice in ice cold PBS. Next, cells were re-suspended in cold lysis buffer containing: 10 mM Tris-HCl, pH 7.4, 10 mM NaCl, 3 mM MgCl2 and 0.1% IGEPAL CA-630(or NP-40). Following centrifugation, supernatant was discarded and cell pellet was used for transposition reaction according to manufacturer’s instructions (Tagment DNA TDE1 Enzyme and Buffer, Illumina 20034210). PCR amplification of transposed DNA, and adaptor incorporation and barcoding was done by KAPA HiFi HotStrat (KK2601, Roche) and Nextera XT index kit (Illumina, 15055293). Libraries were sequenced on an Illumina NovaSeq 6000 sequencer (Illumina, United States) to generate paired-end (100 bp) reads for each sample. RNA-seq read quality was evaluated using FastQC and adapter sequences were removed with Trim Galore. Reads were aligned to the human genome (GRCh38) using bowtie followed by PCR duplicates removal by picard MarkDuplicates. Peak calling was done using MACS2, and peak summits distribution analysis was performed with ‘chipSeeker’ package in R. Coverage around TSS, differential peak analysis and GO term enrichment was performed with ‘recoup’, ’DiffBind’ and ‘clusterProfiler’ packages in R, respectively.

### Toxicity BJAB cells

BJAB cells were used to test potential toxicity of different ASOs using a protocol described in Pollak et al^22^. Shortly, human BJAB cells were cultured in suspension at 37°C in RPMI medium (RPMI, 15% serum, 1% Penicillin-Streptomycin, 1% Glutamine and 1% Sodium Pyruvate). Cells were incubated for 16 hours at 37°c with different ASOs at different concentrations (20nM-5μM), including positive control (ISIS353512), and subsequently harvested. After RNA extraction, human *CCL22* levels were determined with rtPCR, using the following primers- FW- CGCGTGGTGAAACACTTCTA, Rev- GATCGGCACAGATCTCCTTATC.

### ModSpec Analysis

Quantification and analysis was done by Activ Motif, Inc.. Briefly, Samples were analyzed on a triple quadrupole (QqQ) mass spectrometer (Thermo Fisher Scientific TSQ Quantiva) directly coupled with an UltiMate 3000 Dionex nano-liquid chromatography system. Peptides were first loaded onto an in-house packed trapping column (3cm×150µm) and then separated on a New Objectives PicoChip analytical column (10 cm×75 µm). Both columns were packed with New Objectives ProntoSIL C18-AQ, 3µm, 200Å resin. The chromatography gradient was achieved by increasing percentage of buffer B from 0 to 35% at a flow rate of 0.30 µl/min over 45 minutes. Solvent A: 0.1% formic acid in water, and B: 0.1% formic acid in 95% acetonitrile.

The QqQ settings were as follows:

- collision gas pressure of 1.5 mTorr
- Q1 peak width of 0.7 (FWHM)
- cycle time of 2 seconds
- skimmer offset of 10 V
- electrospray voltage of 2.5 kV

Targeted analysis of unmodified and various modified histone peptides was performed. This entire process was repeated three separate times for each sample.

### Asxl3 mice models

Mouse blastocytes’ injection was approved by the Weizmann Institute’s IACUC committee and were carried out in accordance with their approved guidelines.gRNAs were designed using a combination of in-silico design tools, including the MIT CRISPR design tool and sgRNA Designer, Rule set 2 in the Benchling implementations (www.benchling.com).

Genetically modified Asxl-31556* and Asxl3-Ex11 mice were generated at the Transgenic Facility at the Weizmann Institute of Science using CRISPR/Cas9 genome editing in isolated one-cell mouse embryos as described (Gertsenstein and Nutter 2021). C57Bl/6JOlaHsd mice were purchased from Envigo, Israel and maintained in specific pathogen-free (SPF) conditions. Mice were maintained on a 12 hr light/dark cycle, and food and water were provided ad libitum. Cas9-gRNA ribonucleoproteins (RNP) complexes together with donor repair template were delivered to one-cell embryos via electroporation, using the Biorad Genepulser. Electroporated embryos were transferred into the oviducts of pseudopregnant ICR females (Envigo, Israel).

Asxl3-1556* was established using a single gRNA that was designed to target gene Asxl3 (5’-TGTAGCAATCAGTATAACCC -3’). A single-stranded oligodeoxynucleotide (ssODN) donor repair template flanked asymmetrically by homology arms to each of the 5′ and 3′ insertion sites was designed (TGAGCAAGCACGAAACTCAAGTACTTTCAAGAAAGAAACAGACACAGCCTGTAGCAATCAaTAcAAtCCtGGAAAtCGCATaTGCTaGAGTGAAGACCCAATGAGAAACACAGCACCGCCTGTGGTTAGTCATTCAAGCTCAAGTAA). Cas9 nuclease, crRNA, tracrRNAs and ssODN were purchased from Integrated DNA Technologies (IDT).

Mice were genotyped using the sequencing forward primer (CAAGTGAAGGGCTTGACCAT), and two reverse primers (WT – TGCGGTTTCCTGGGTTATAC, Mutant – ATGCGATTTCCAGGATTGTAT)

Two gRNAs were designed to target Exon 11 of gene Asxl3 (5’-TGTCGGCTTATTTCCTAGAT-3’) and (5’-CTAGCATGTCACATAATAAC-3’)) respectively. Cas9 nuclease, crRNA, tracrRNAs and ssODN were purchased from Integrated DNA Technologies (IDT).

Mice were genotyped using the sequencing forward primer (CACAATTACAGGCAGGGACAG), and reverse primer (ACTCTAGAATACCCCAGCTGC).

Subsequent animal experiments described herein were approved by the Sheba Medical Center IACUC (1293/21/ANIM), and the colony was maintained in the Sheba animal facility. Mice were genotyped using the sequencing forward primer (CAAGTGAAGGGCTTGACCAT), and two reverse primers (WT – TGCGGTTTCCTGGGTTATAC, Mutant – ATGCGATTTCCAGGATTGTAT). All behavioral assays (as described is **Figure S7B**) were performed in MD biosciences CRO.

### Statistical analysis

Unless stated, each experiment was performed at least three times for each condition. Statistical analyses were performed with R or Prism v11 (GraphPad) and statistical tests are noted in figure legends.

## List of Supplementary materials

Figures S1 to S8

Tables S1 to S2

## Data Availability

All data produced in the present study are available upon reasonable request to the authors

## Acknowledgements

The authors would like to thank Dr. Daoud Sheban, Dr. Anat Gaoni-Yogev for valuable comments and discussions. NM is a schooler of the Nehemia Rubin Excellence in Biomedical Research, the TELEM Program at the Sheba medical Center. The work was supported by the Lowy Medical Research Initiative.

## Author contributions

Conceptualization :NM, IS, DD, GH, GR

Methodology :IS, SR, NBH, TM, SA, YR, SAN, AY, ESG

Investigation :NM, IS, SR, NBH, TM, SA, AY, ESG, YR, RHK, SBD

Formal Analysis :ON, SBD

Writing – Original Draft :NM, IS, DD

Supervision :NM, GR, DD

Funding Acquisition :DD, NM

## Competing interests

The authors declare no competing interests.

## Data and materials availability

All RNA-seq, Methylation and ATAC-seq data are deposited in Zenodo, accession number TBD.

**Supplementary Table 1 –** Full list of primers used in this study.

**Supplementary Table** 2 – Full list of ASOs used in this study.

---

**Figure S1.**
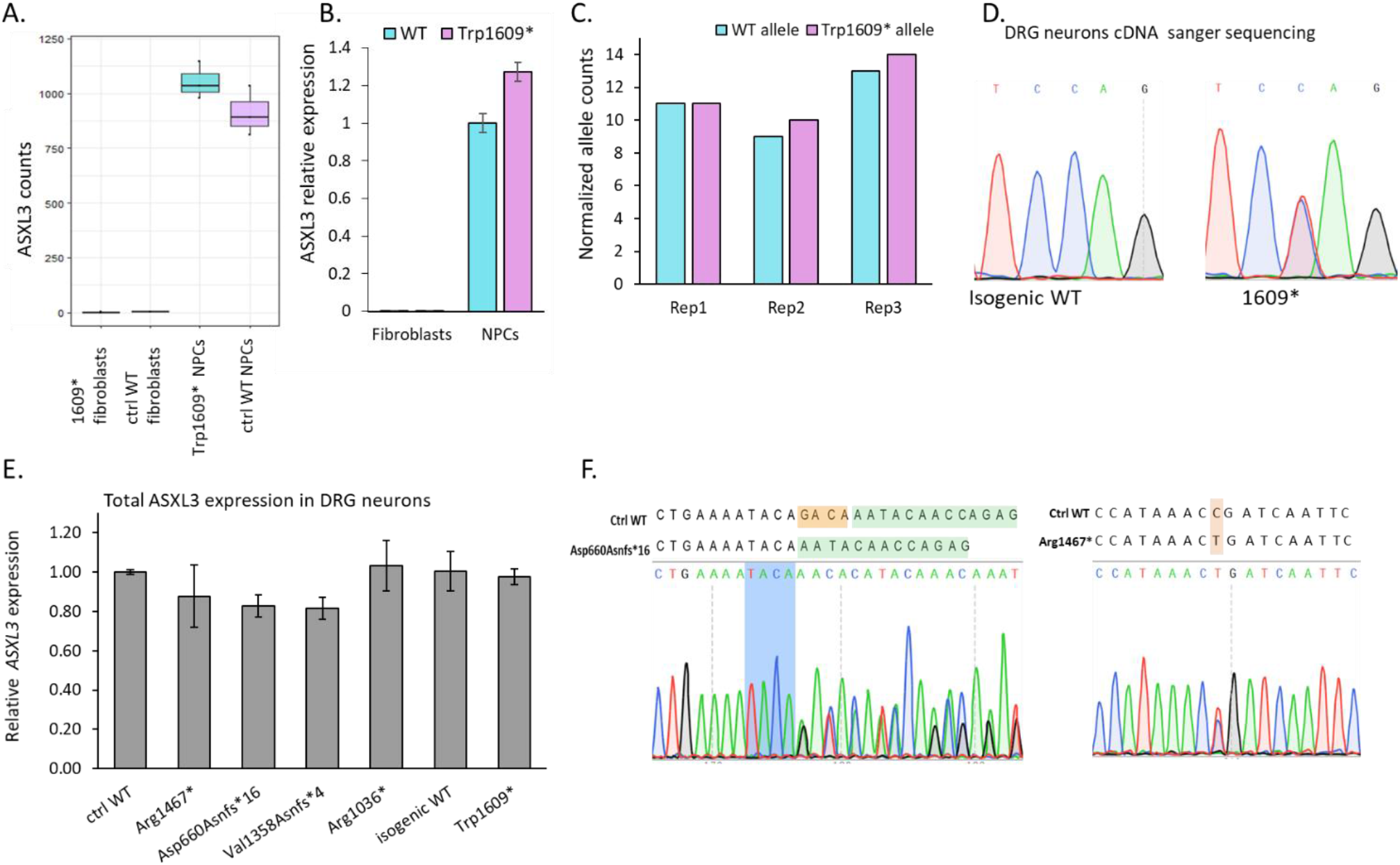
Molecular characterization of ASXL3 in BRS patient-derived cellular models. (A) *ASXL3* read counts as seen in RNAseq of Trp1609* and isogenic WT -derived NPCs and fibroblasts. (B) Transcript level of *ASXL3* using RT-qPCR on patient and isogenic WT -derived NPCs and fibroblasts (normalized to GAPDH, n=3). (C) wt and mutant allele counts measured by RNA-seq in patient -derived NPCs. (D) Sanger sequencing of Trp1609* variant region in patient and isogenic WT -derived DRG neurons. (E) Transcript level of *ASXL3* using RT-qPCR in patient and control– derived DRG neurons. (normalized to GAPDH, n=3). (F) Sanger sequencing of variant region in Asp660Asnfs*16 (primers 23+24 in Table 1) and Arg1467* (primers 31+32 in Table 1) -derived DRG neurons.

**Figure S2.**
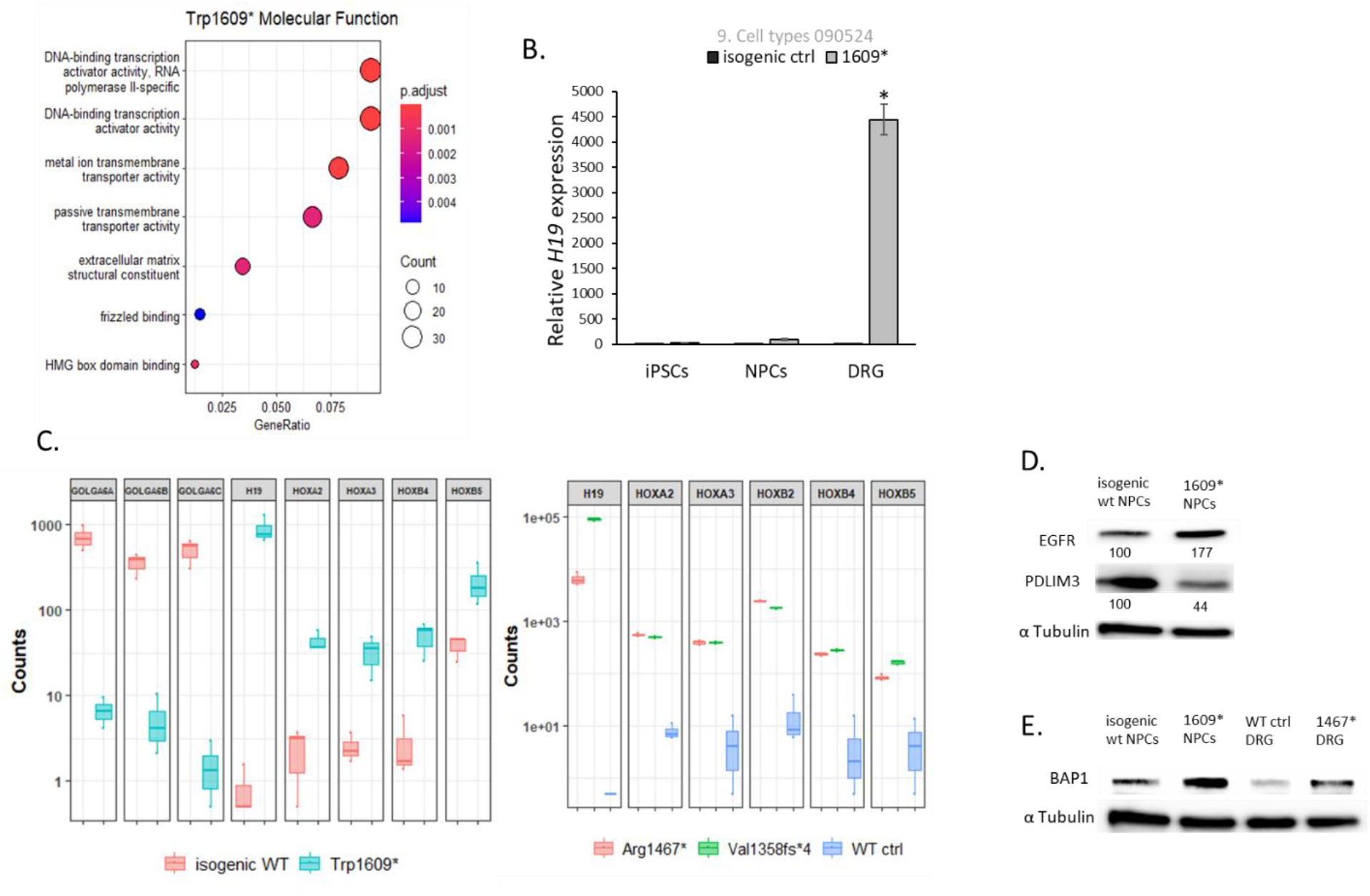
Molecular phenotype characterization of patients-derived cellular models. (A) GO-enrichment of molecular function for differentially expressed genes in patient –derived DRG neurons. (B) Transcript level of *H19* using RT-qPCR in patient and control –derived DRG neurons, NPCs and iPSCs. (normalized to GAPDH, n=3, * p value<0.01. (C) Differentially expressed gene counts as seen in RNAseq in patient-derived cell lines and control (isogenic control and NHD), all differentiated to DRG neurons (D) Western blot of PDLIM3 and EGFR differentially expressed proteins in patient and control –derived NPCs. α-Tubulin was used as loading control. Band intensity was measured using ImageLab software and normalized to WT control. (E) Western blot of BAP1 protein in patient and control –derived NPCs and DRG neurons. α-Tubulin was used as loading control.

**Figure S3.**
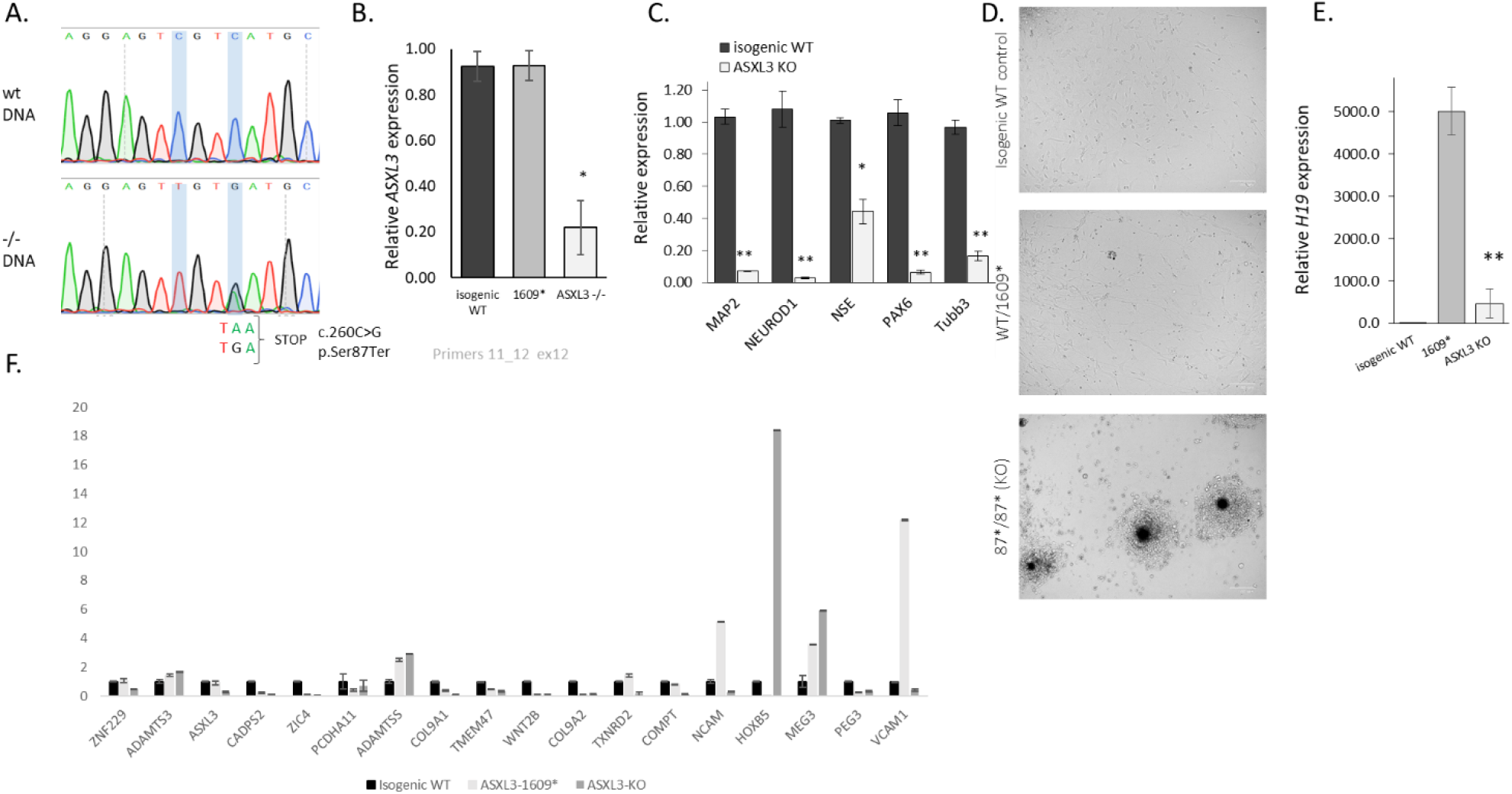
ASXL3 bi-allelic KO affect neuronal differentiation. (A) Sanger sequencing of the incorporated premature termination codon in ASXL3-KO and control – derived DRG neurons. (B) Transcript level of *ASXL3* using rRT-qPCR on ASXL3-KO, patient and isogenic WT -derived DRG neurons (normalized to GAPDH, n=3). (C) Transcript level of differentiation markers in isogenic WT vs. ASXL3-KO -derived DRG neurons, * p-value< 0.01, ** p-value< 0.001 (normalized to GAPDH, n=3). (D) Bright-field image of ASXL3-KO, patient and isogenic WT –derived DRG neurons during differentiation process at day 7. (E) Transcript level of *H19* using rRT-qPCR on ASXL3-KO, patient and isogenic WT –derived DRG neurons, ** p-value< 0.001(normalized to GAPDH, n=3).

**Figure S4.**
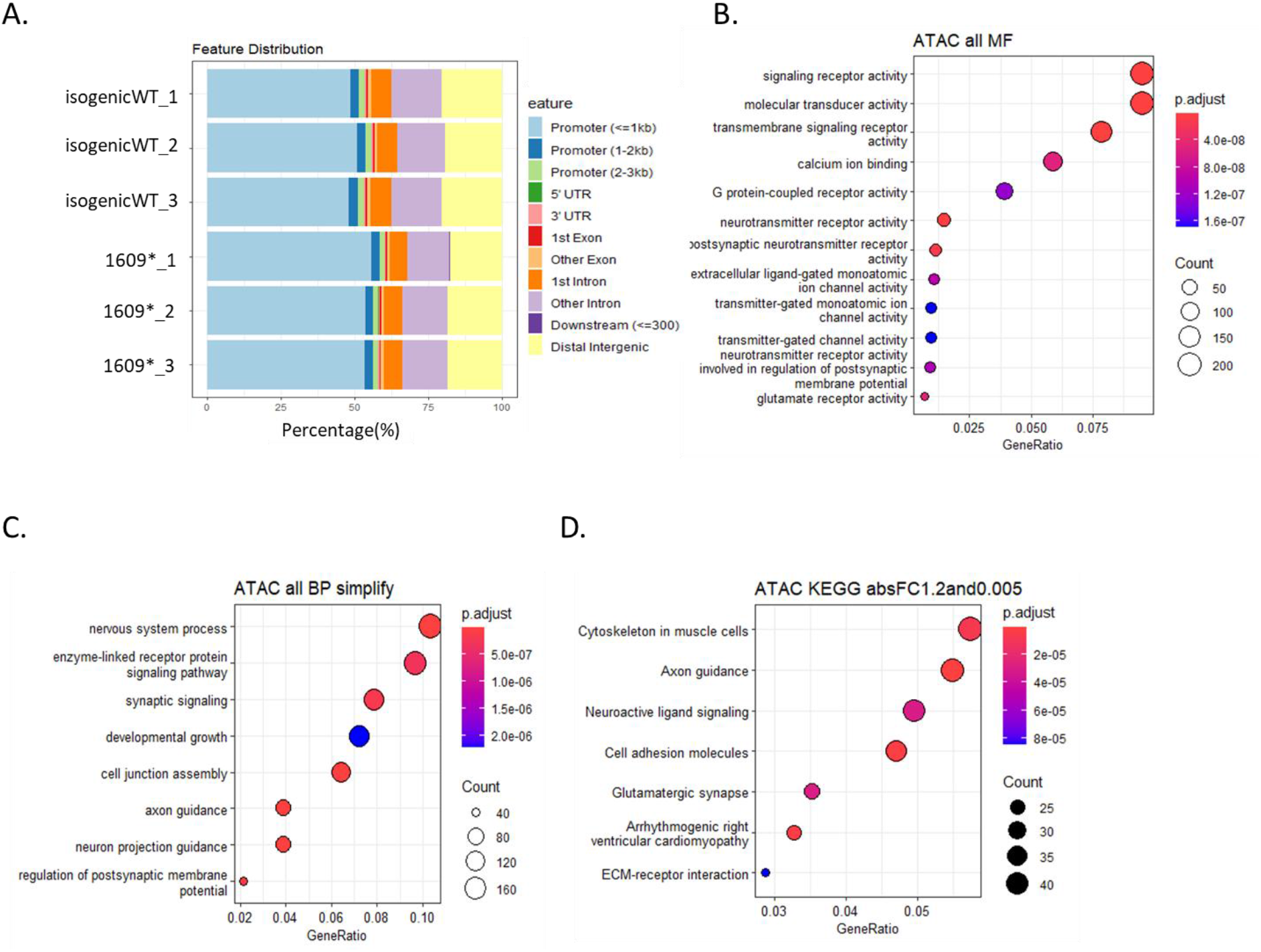
Chromatin accessibility and epigenetic signature in cellular models. (A) Accessible regions in the chromatin as identified in ATAC-seq on patient-derived and isogenic control DRG (B) GO-enrichment of molecular function for differentially enriched regions in patient – derived DRG neurons. (C) GO-enrichment of Biological Processes for differentially enriched regions in patient –derived DRG neurons. (D) Pathway analysis (KEGG) for differentially enriched regions in patient –derived DRG neurons.

**Figure S5.**
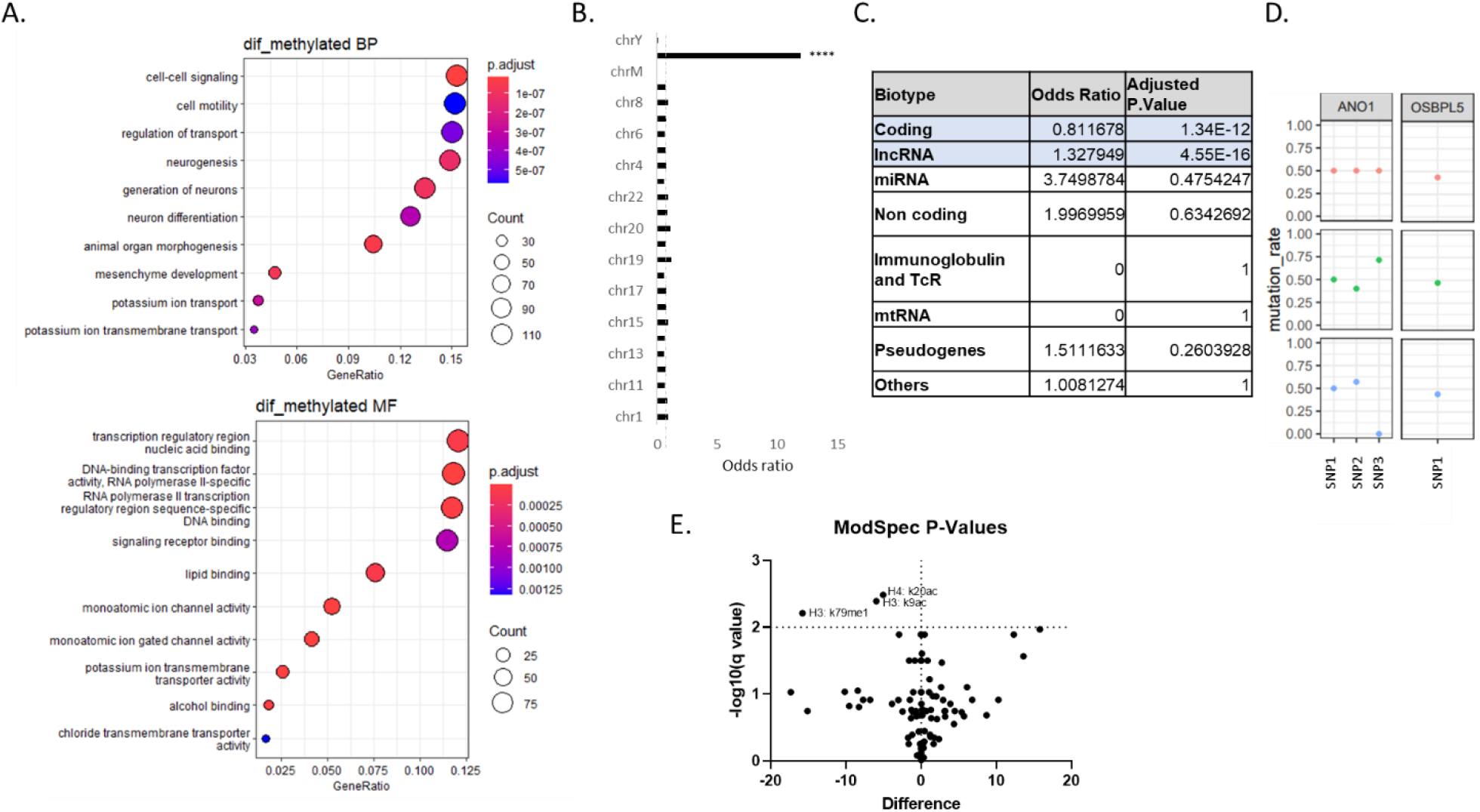
Differential methylation in ASXL3-1609* DRG neurons comparing to isogenic control. (A) GO Enrichment for differentially methylated regions between Patient-derived DRG neurons (ASXL3-1609*) and isogenic control. (B) chromosomal mapping of differentially methylated regions shows a non-random distribution across the genome, with a notable enrichment on chromosome X. (C) Differentially methylated regions analysis based on transcript type shows a significant enrichment of lncRNA. (D) Bi-allelic expression of representative imprinted genes seen in heterozygote SNPs in patient-derived DRG neurons (ASXL3-1609*). Heterozygote SNPs were identified based on WGS data, and transcripts’ levels were analyzed in polyA RNA-seq data (n=3). (E) ModSpec analysis of histone modification in patient-derived DRG neurons and their isogenic control (n=3).

**Figure S6.**
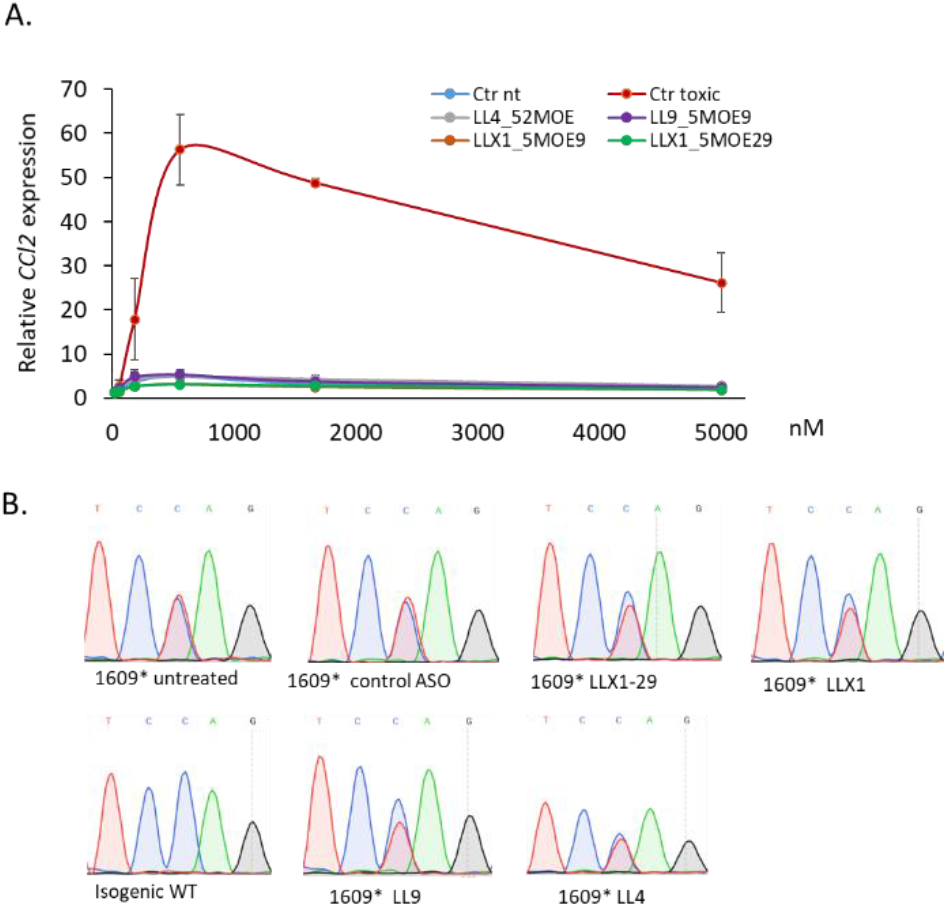
ASO toxicity prediction assay and selectivity efficiency. (A) Toxicity prediction using rtPCR of *CCL2* in BJAB cells treated with increased concentrations of ASO. ASO ISIS353512 was used as a toxic control (red), and ASO ISIS104838 was used as non-toxic control. CCL2 response was calculated as the ration of CCL2 level with tested ASO relative to untreated cells (n=3) (B) Sanger sequencing of isogenic control and patient –derived DRG neurons following ASO treatment (5uM) for 72 hours.

**Figure S7:**
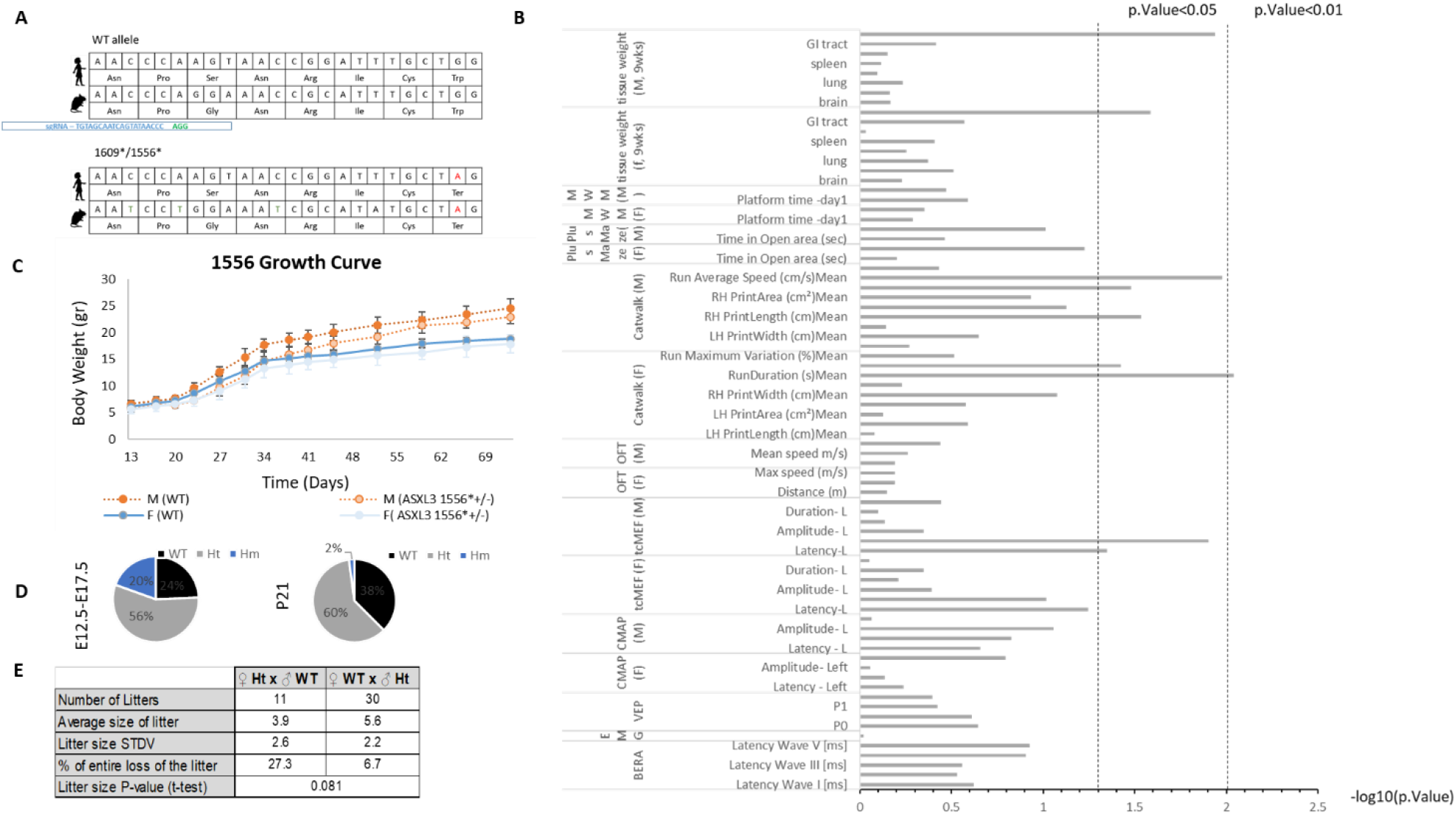
Asxl3-1556* mouse model shows mild growing retardation and a significant shift in mendelian ratios. (A) Targeting scheme for a CRISPR/Cas9 zygote injection, aiming to induce a specific mutation in mAsxl3, recapitulating the patient’s mutation. (B) Asxl3-155* heterozygote mice and WT littermates were subjected to multiple standardized assays assessing motor function, sensory response, cognitive function and electrophysiological parameters. Tests were conducted at two time points (weeks and 9 weeks), apart fro electrophysiological tests which were conducted only at the 9-weeks time point. Following these tests, the mice were euthanized and exploratory anatomical estimation was carried out to assess gross malformations. P.value was calculated for all results, showing no phenotype or a mild effect. The table summarizes results from male mice, at 9 weeks (n=5 per group). (C) Growth curves for Asxl3^1556/+^ heterozygote and WT littermates, weekly weightings from day13 to 70 . Male (N>8, student’ t test, p value<0.0005 for all time-points, except for days 59 and 66 and Females (N>4, student’ t test, not significant). (D) Genotyping of Asxl3-1556 embryos and neonates (n=10 litters, 82 pups) show normal mendelian distribution, non-consistent with the lack of homozygote pups>P2 (n=48, χ2=0.00085). (E) Maternal care follow-up for Asxl3^1556/+^ heterozygote females and wildtype littermates, show less litters for HT females, and smaller litter size at weaning.

**Figure S8.**
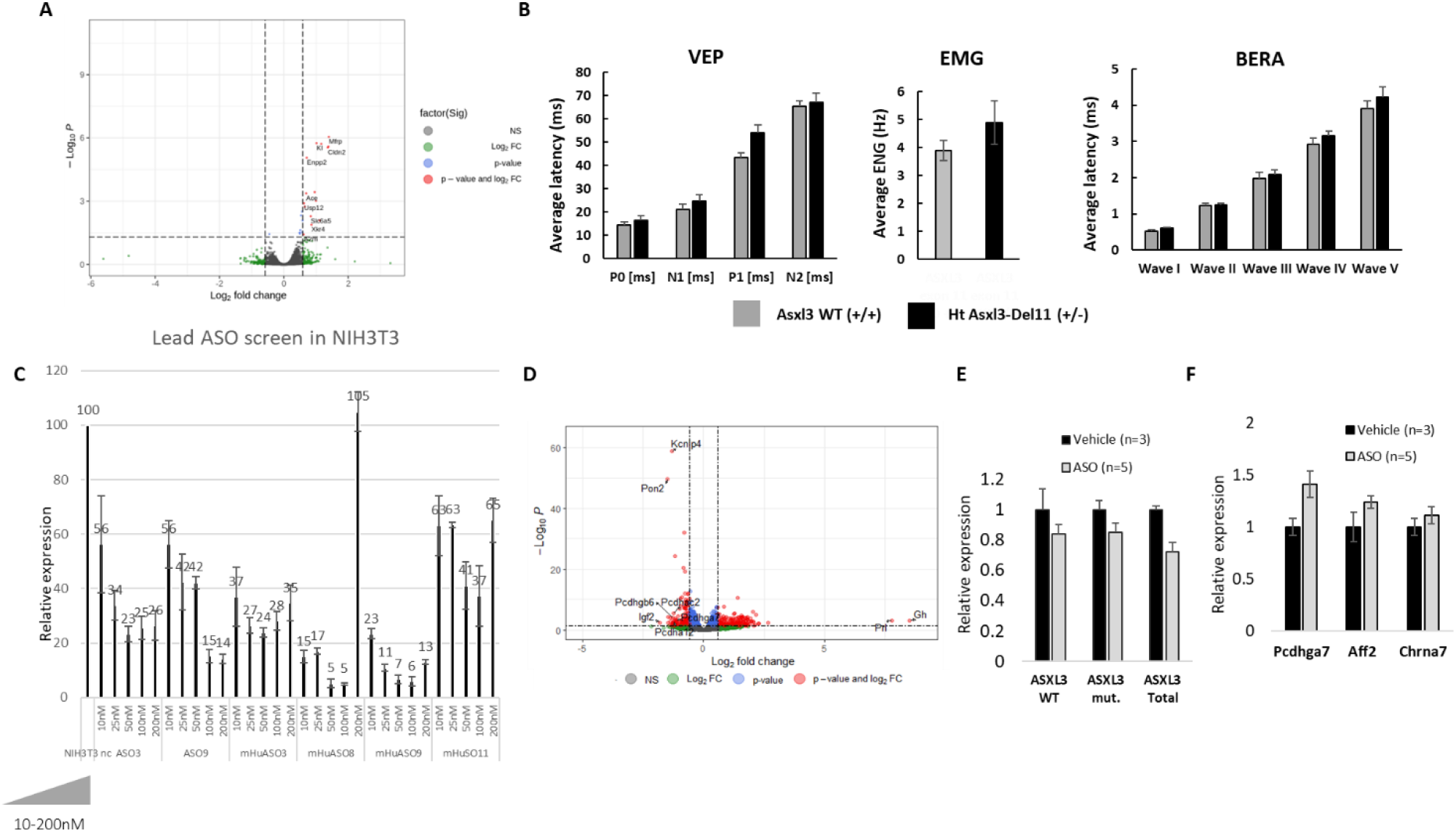
Partial reduction of Asxl3 in mice brain, by genetic manipulation of ASO injection, is tolerated. (A) Volcano plot of the results of RNA-seq-based differential gene expression analysis between heterozygote Asxl3-Del11 (+/-) and WT littermates, showing minimal differentially expressed genes (n=3 per group). (B) Visual evoked potentials (VEP), frequency of spontaneous action potentials (electromyography, EMG), and brainstem auditory evoked response audiometry (BERA) were recorded in heterozygous Asxl3-Del11 (+/−) mice and WT littermates at 9 weeks of age (n=5 per group). No statistically significant differences were observed between genotypes across all measurements. (C) Asxl3 transcript levels in 3T3 cells, as measured by rt-qPCR following ASO treatments. Cells were transfected with different concentrations of different ASO, in accordance to the bar plot. Asxl3 levels were quantified after 48 hours of treatment, and normalized to a house keeping gene. (D) Volcano plot of the results of RNA-seq-based differential gene expression analysis between WT mice injected with PBS or Asxl3-targeting ASO, ten days post injection (n=3 per group). (E) Asxl3-1556* mice were injected with Asxl3-targeting ASO (6mg/kg) or Vehicle. Study was terminated 28-days post injection, and brain tissue from the injected mice was analyzed for Asxl3 levels (total, WT allele and mutated allele), showing 20-30% reduction in Asxl3 expression, with bi-allelic contribution. (F) Genes that were highlighted as differentially expressed following Asxl3-targeting ASO treatment in WT mice (ass seen in **Figure S8D**) were evaluated by rt-qPCR following ASO treatment (28 days) in Asxl3-1556* mice, showing an increase in these mice as well.

## Notes

### Competing Interest Statement

The authors have declared no competing interest.

### Author Declarations

Human subjects research was conducted in accordance with protocols approved by the Sheba Medical Center Institutional Review Board.

