## Supplemental Data for "Truncated ASXL3 Alters Chromatin Accessibility and Epigenetic Landscape in Bainbridge-Ropers Syndrome Suggesting a Gain-of-Function Etiology"

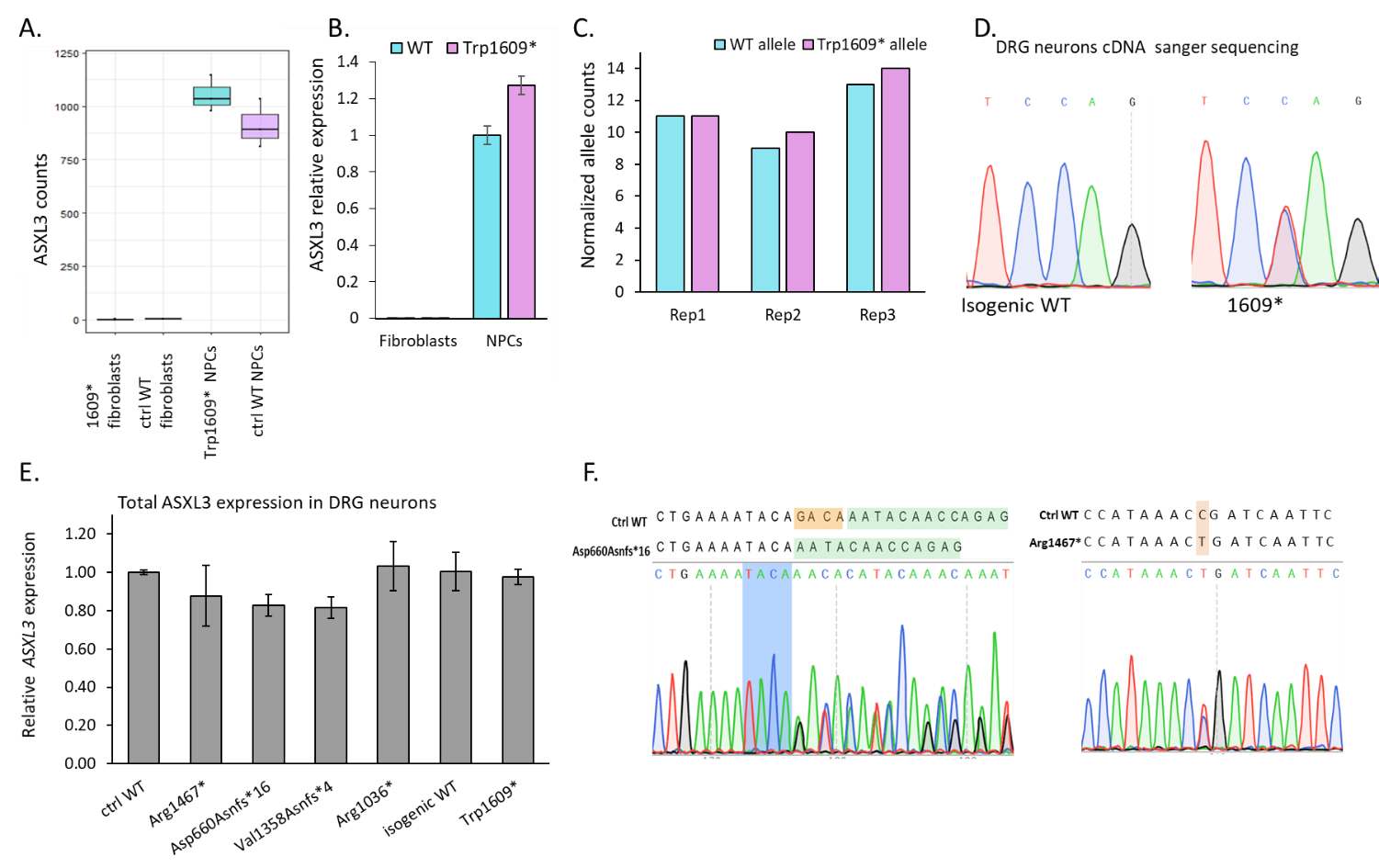

***Figure 1S. Molecular characterization of ASXL3 in BRS patient-derived cellular models.***

(A) *ASXL3* read counts as seen in RNAseq of Trp1609* and isogenic WT -derived NPCs and fibroblasts. (B) Transcript level of *ASXL3* using RT-qPCR on patient and isogenic WT -derived NPCs and fibroblasts (normalized to GAPDH, n=3). (C) wt and mutant allele counts measured by RNA-seq in patient -derived NPCs. (D) Sanger sequencing of Trp1609* variant region in patient and isogenic WT -derived DRG neurons. (E) Transcript level of *ASXL3* using RT-qPCR in patient and control–derived DRG neurons. (normalized to GAPDH, n=3). (F) Sanger sequencing of variant region in Asp660Asnfs*16 (primers 23+24 in Table 1) and Arg1467* (primers 31+32 in Table 1) -derived DRG neurons.

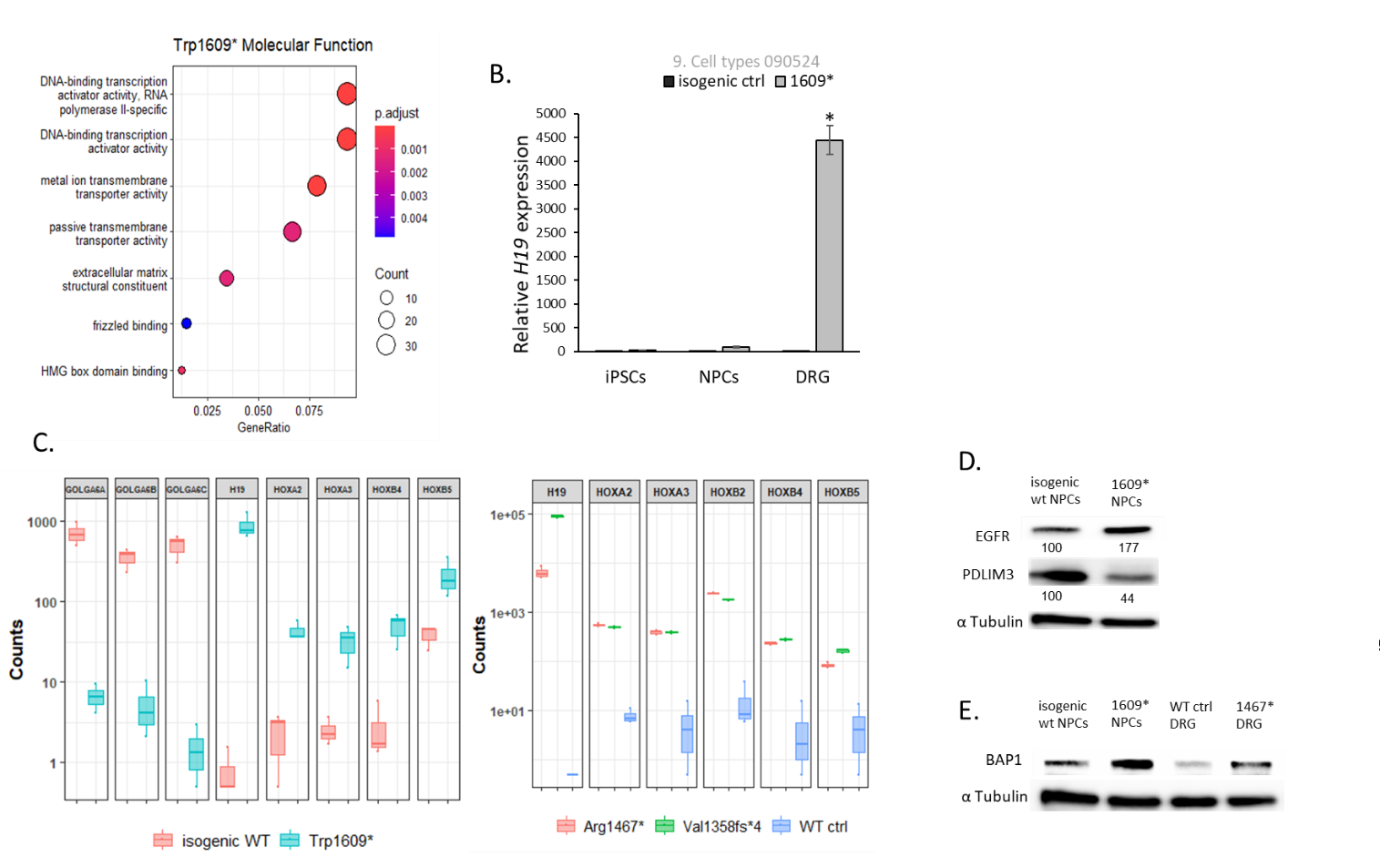

***Figure 2S. Molecular phenotype characterization of patients-derived cellular models.***

(A) GO-enrichment of molecular function for differentially expressed genes in patient –derived DRG neurons. (B) Transcript level of *H19* using RT-qPCR in patient and control –derived DRG neurons, NPCs and iPSCs. (normalized to GAPDH, n=3, * p value<0.01. (C) Differentially expressed gene counts as seen in RNAseq in patient-derived cell lines and control (isogenic control and NHD), all differentiated to DRG neurons (D) Western blot of PDLIM3 and EGFR differentially expressed proteins in patient and control –derived NPCs. α-Tubulin was used as loading control. Band intensity was measured using ImageLab software and normalized to WT control. (E) Western blot of BAP1 protein in patient and control –derived NPCs and DRG neurons. α-Tubulin was used as loading control.

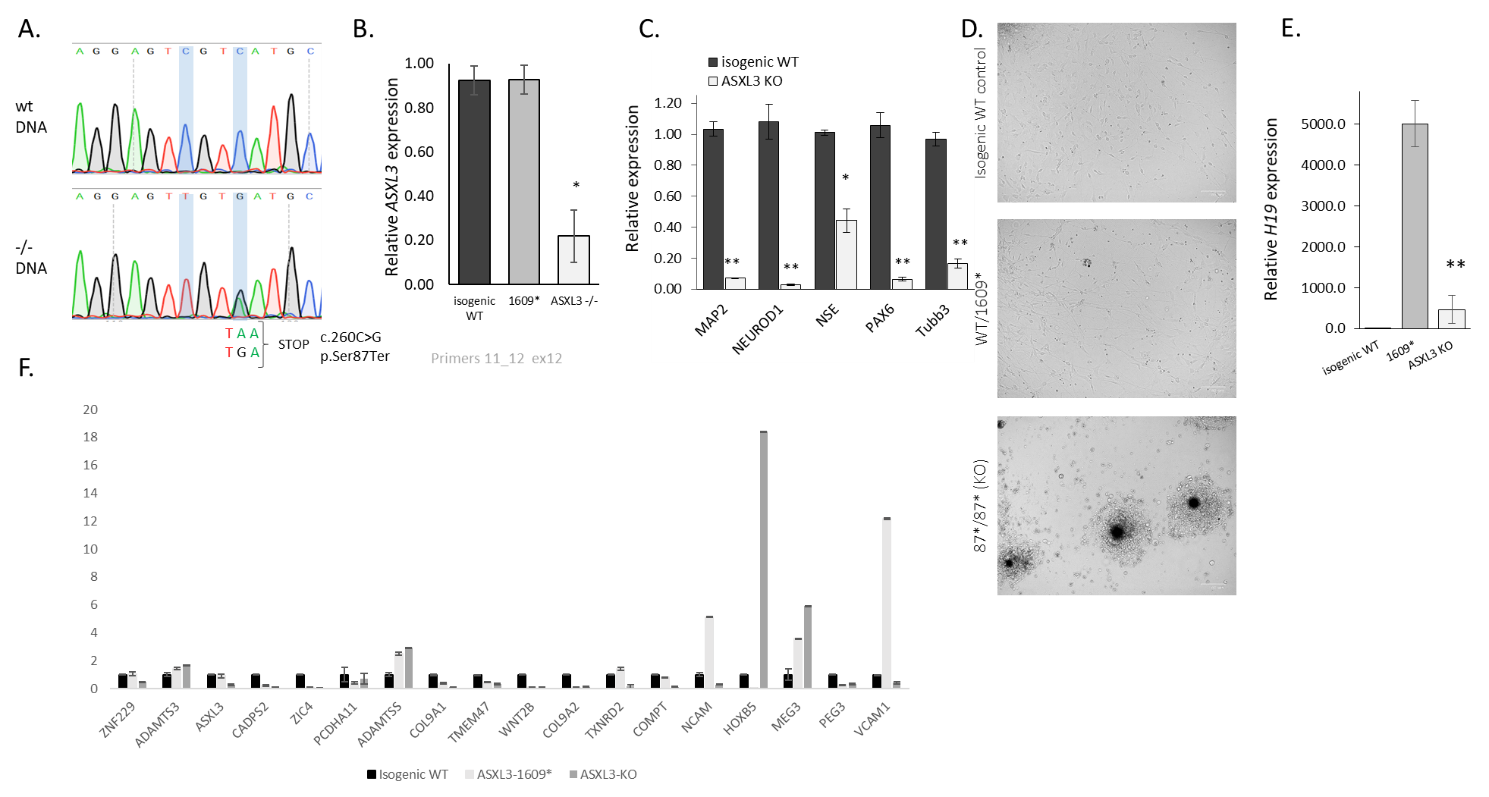

***Figure 3S. ASXL3 bi-allelic KO affect neuronal differentiation.***

(A) Sanger sequencing of the incorporated premature termination codon in ASXL3-KO and control –derived DRG neurons. (B) Transcript level of *ASXL3* using rRT-qPCR on ASXL3-KO, patient and isogenic WT -derived DRG neurons (normalized to GAPDH, n=3). (C) Transcript level of differentiation markers in isogenic WT vs. ASXL3-KO -derived DRG neurons, * p-value< 0.01, ** p-value< 0.001 (normalized to GAPDH, n=3). (D) Bright-field image of ASXL3-KO, patient and isogenic WT –derived DRG neurons during differentiation process at day 7. (E) Transcript level of *H19* using rRT-qPCR on ASXL3-KO, patient and isogenic WT –derived DRG neurons , ** p-value< 0.001(normalized to GAPDH, n=3).

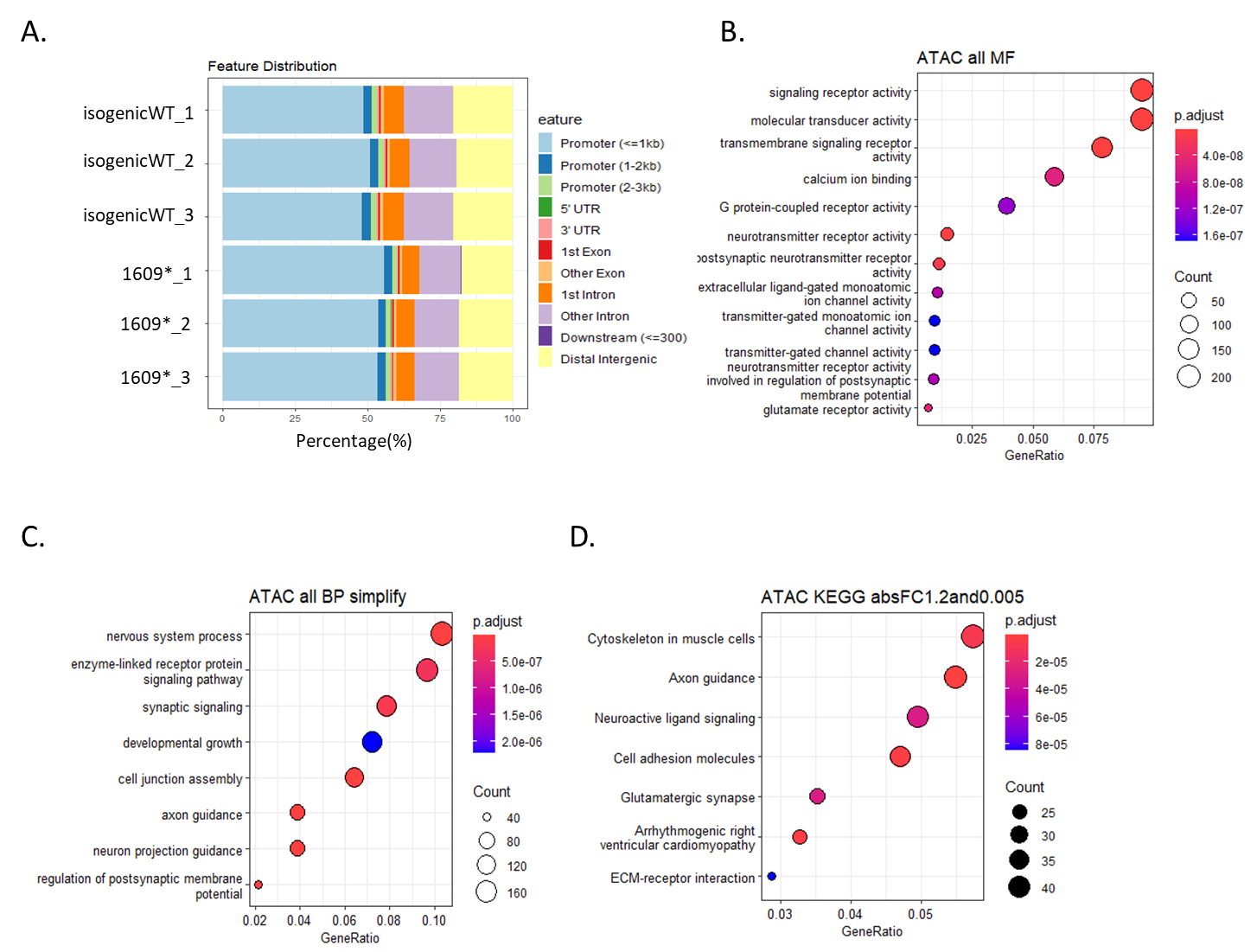

***Figure 4S. Chromatin accessibility and epigenetic signature in cellular models.***

(A) Accessible regions in the chromatin as identified in ATAC-seq on patient-derived and isogenic control DRG (B) GO-enrichment of molecular function for differentially enriched regions in patient –derived DRG neurons. (C) GO-enrichment of Biological Processes for differentially enriched regions in patient –derived DRG neurons. (D) Pathway analysis (KEGG) for differentially enriched regions in patient –derived DRG neurons.

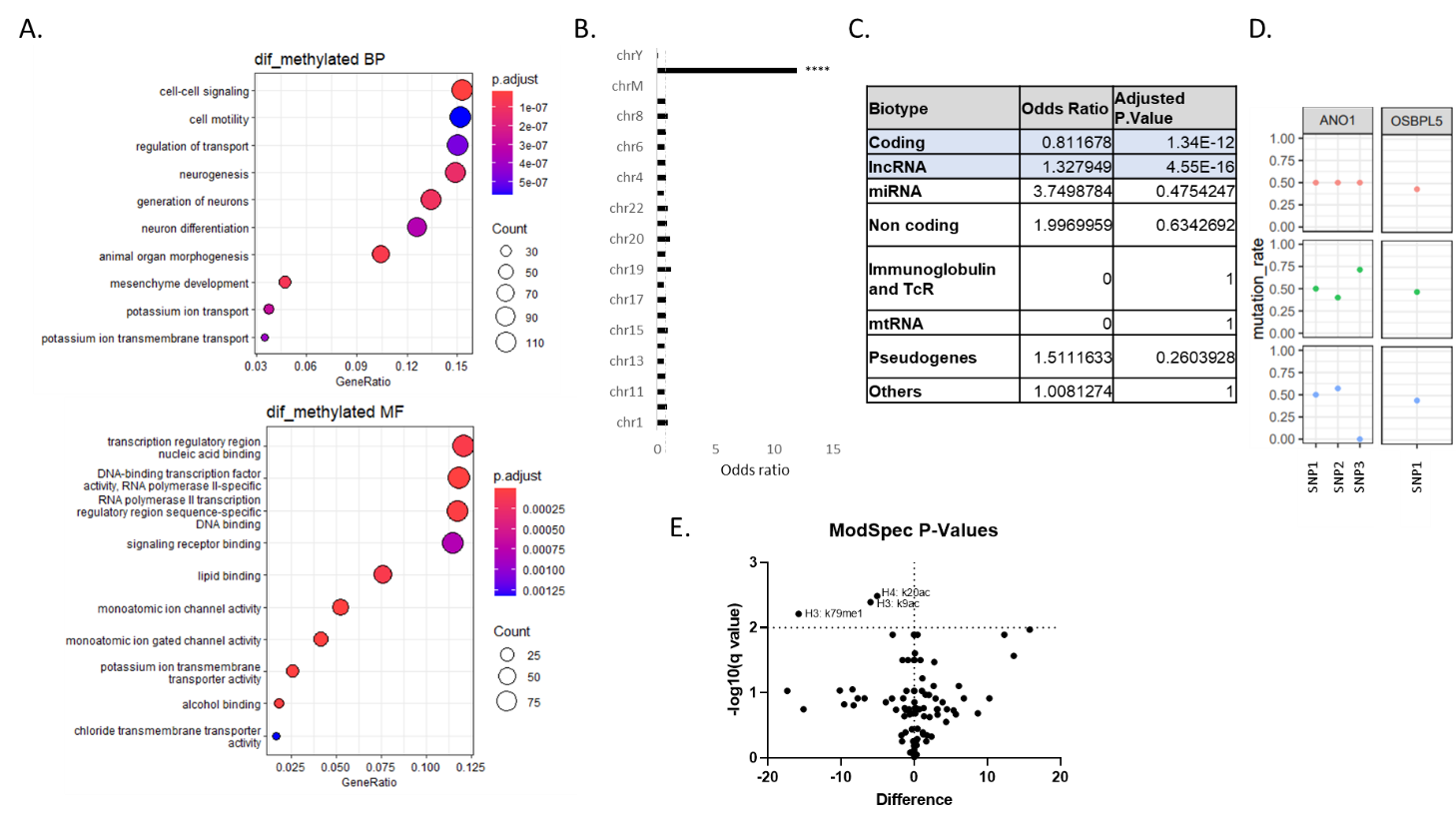

***Figure 5S. Differential methylation in ASXL3-1609* DRG neurons comparing to isogenic control***

(A) GO Enrichment for differentially methylated regions between Patient-derived DRG neurons (ASXL3-1609*) and isogenic control. (B) chromosomal mapping of differentially methylated regions shows a non-random distribution across the genome, with a notable enrichment on chromosome X. (C) Differentially methylated regions analysis based on transcript type shows a significant enrichment of lncRNA. (D) Bi-allelic expression of representative imprinted genes seen in heterozygote SNPs in patient-derived DRG neurons (ASXL3-1609*). Heterozygote SNPs were identified based on WGS data, and transcripts’ levels were analyzed in polyA RNA-seq data (n=3). (E) ModSpec analysis of histone modification in patient-derived DRG neurons and their isogenic control (n=3).

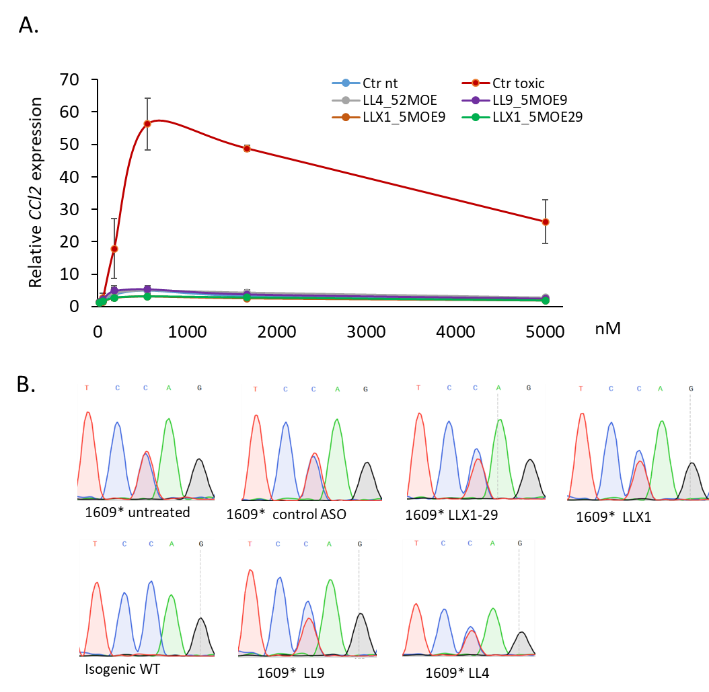

***Figure 6S. ASO toxicity prediction assay and selectivity efficiency.***

(A) Toxicity prediction using rtPCR of *CCL2* in BJAB cells treated with increased concentrations of ASO. ASO ISIS353512 was used as a toxic control (red), and ASO ISIS104838 was used as non-toxic control. CCL2 response was calculated as the ration of CCL2 level with tested ASO relative to untreated cells (n=3) (B) Sanger sequencing of isogenic control and patient –derived DRG neurons following ASO treatment (5uM) for 72 hours.

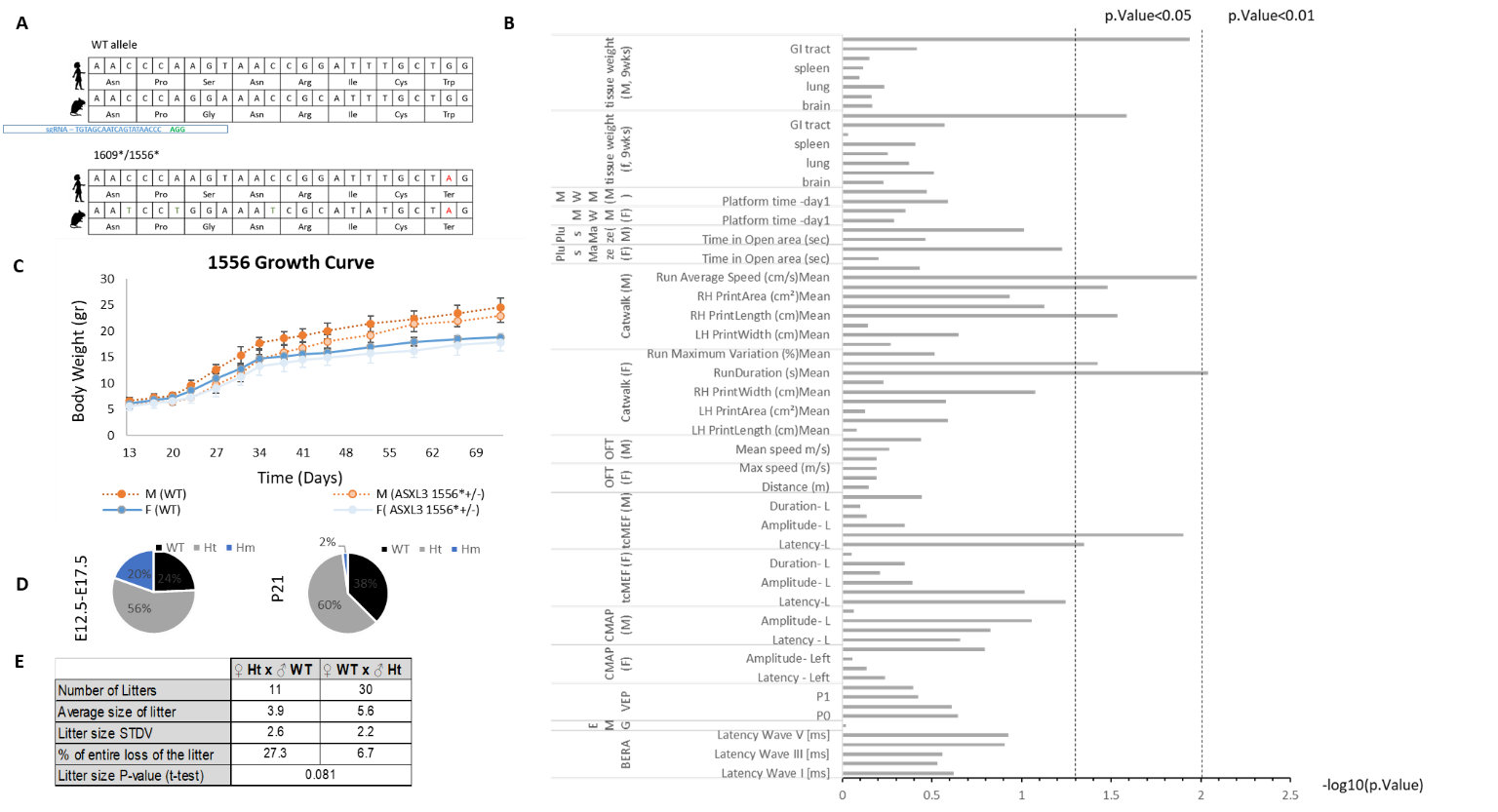

***Figure 7s: Asxl3-1556* mouse model shows mild growing retardation and a significant shift in mendelian ratios.***

(A) Targeting scheme for a CRISPR/Cas9 zygote injection, aiming to induce a specific mutation in mAsxl3, recapitulating the patient’s mutation. (B) Asxl3-155* heterozygote mice and WT littermates were subjected to multiple standardized assays assessing motor function, sensory response, cognitive function and electrophysiological parameters. Tests were conducted at two time points ( weeks and 9 weeks), apart fro electrophysiological tests which were conducted only at the 9-weeks time point. Following these tests, the mice were euthanized and exploratory anatomical estimation was carried out to assess gross malformations. P.value was calculated for all results, showing no phenotype or a mild effect. The table summarizes results from male mice, at 9 weeks (n=5 per group). (C) Growth curves for Asxl3^1556/+^ heterozygote and WT littermates, weekly weightings from day13 to 70 . Male (N>8, student’ t test, p value<0.0005 for all time-points, except for days 59 and 66 and Females (N>4, student’ t test, not significant). (D) Genotyping of Asxl3-1556 embryos and neonates (n=10 litters, 82 pups) show normal mendelian distribution, non-consistent with the lack of homozygote pups>P2 (n=48, χ2=0.00085). (E) Maternal care follow-up for Asxl3^1556/+^ heterozygote females and wildtype littermates, show less litters for HT females, and smaller litter size at weaning.

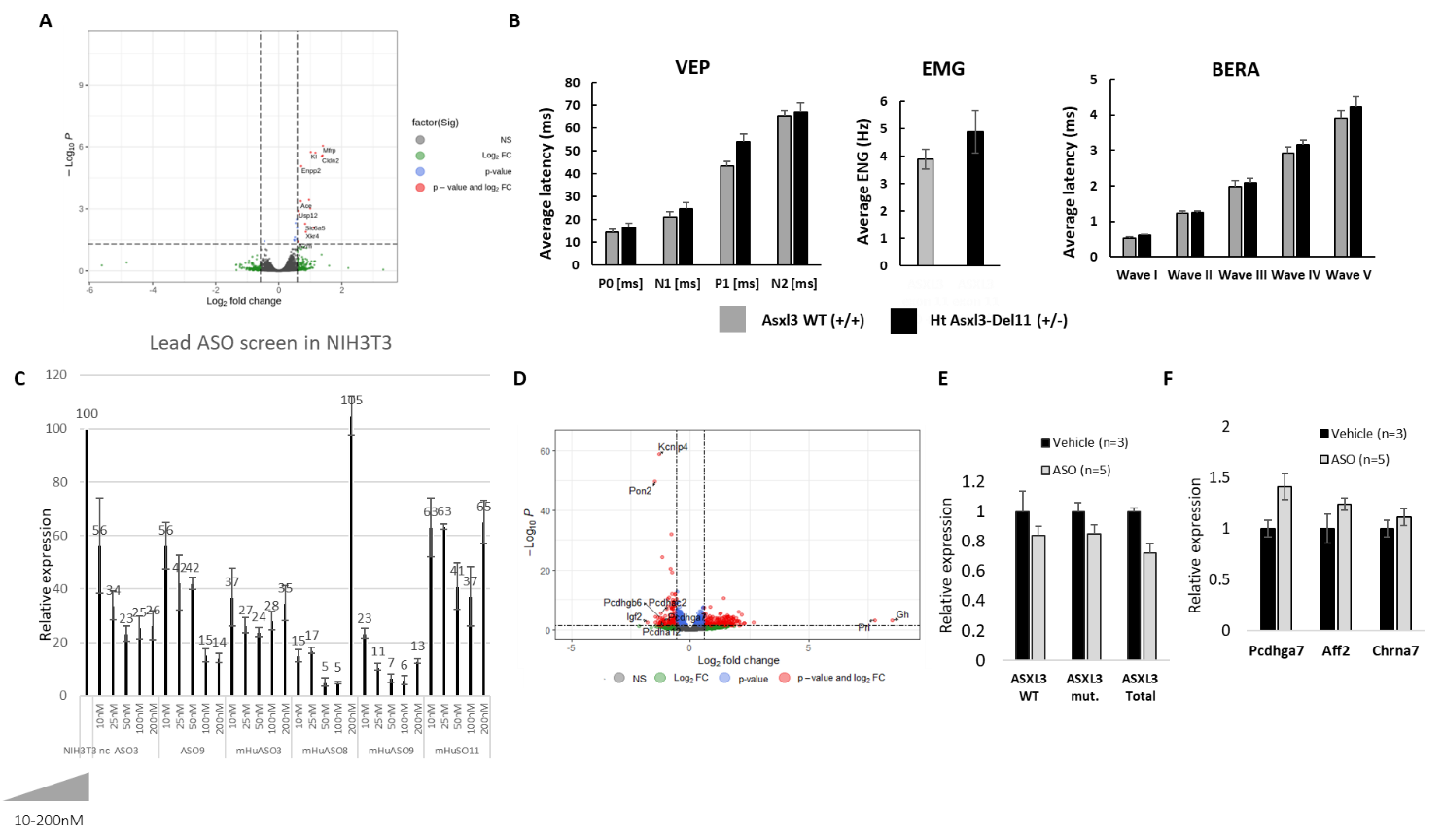

***Figure 8S – Partial reduction of Asxl3 in mice brain, by genetic manipulation of ASO injection, is tolerated****.* (A) Volcano plot of the results of RNA-seq-based differential gene expression analysis between heterozygote Asxl3-Del11 (+/-) and WT littermates, showing minimal differentially expressed genes (n=3 per group). (B) Visual evoked potentials (VEP), frequency of spontaneous action potentials (electromyography, EMG), and brainstem auditory evoked response audiometry (BERA) were recorded in heterozygous Asxl3-Del11 (+/−) mice and WT littermates at 9 weeks of age (n=5 per group). No statistically significant differences were observed between genotypes across all measurements. (C) Asxl3 transcript levels in 3T3 cells, as measured by rt-qPCR following ASO treatments. Cells were transfected with different concentrations of different ASO, in accordance to the bar plot. Asxl3 levels were quantified after 48 hours of treatment, and normalized to a house keeping gene. (D) Volcano plot of the results of RNA-seq-based differential gene expression analysis between WT mice injected with PBS or Asxl3-targeting ASO, ten days post injection (n=3 per group). (E) Asxl3-1556* mice were injected with Asxl3-targeting ASO (6mg/kg) or Vehicle. Study was terminated 28-days post injection, and brain tissue from the injected mice was analyzed for Asxl3 levels (total, WT allele and mutated allele), showing 20-30% reduction in Asxl3 expression, with bi-allelic contribution. (F) Genes that were highlighted as differentially expressed following Asxl3-targeting ASO treatment in WT mice (ass seen in **Figure S8D**) were evaluated by rt-qPCR following ASO treatment (28 days) in Asxl3-1556* mice, showing an increase in these mice as well.

**Supplementary Table 1 –** Full list of primers used in this study.

**Supplementary Table 2** – Full list of ASOs used in this study.
